# Deep Learning Identified Extra-Prostatic Extension and Seminal Vesicle Invasion as an MRI Biomarker for Prostate Cancer Outcomes

**DOI:** 10.1101/2024.12.31.24319822

**Authors:** Sajid Hossain, Saeed Hossain, Durga Sritharan, Daniel Fu, Aishwarya Nene, Jahid Hossain, Saahil Chadha, Issac Kim, MingDe Lin, Mariam Aboian, Sanjay Aneja

## Abstract

Current risk stratification methods for localized prostate cancer (PCa) are reliant on clinical and pathological variables that do not easily account for location of cancer spread. Prostate MRI is a helpful tool to identify anatomic extra-prostatic cancer spread (EPE) and seminal vesicle invasion (SVI) but is subject to radiologist expertise and inter-observer variation. We report deep learning models which provide objective end-to-end evaluation of EPE and SVI on prostate MRI. Both EPE and SVI models demonstrate high discriminatory ability on three held-out test sets spanning different clinical settings, equipment manufacturers, and MRI magnet strengths. Interpretability studies suggest both EPE and SVI models identify clinically-relevant anatomic regions. Lastly, we show that classification of EPE and SVI by our models is independently associated with increased risk of biochemical recurrence (BCR) following localized treatment. Furthermore, we demonstrate that our models can be easily integrated to well-established risk stratification methods (NCCN and UCSF-CAPRA) for improved ability to identify high risk PCa phenotypes.

## Introduction

Prostate cancer (PCa) is the most common cause of non-cutaneous cancer among men worldwide, with an estimated 1.4 million new cases and 375,000 deaths annually.^1^ Over the past decade, a number of novel agents have been developed to be used in combination with localized treatment with hopes of optimizing patient outcomes. While radiation therapy (RT) and radical prostatectomy (RP) remain the mainstays of definitive treatment for localized PCa, the use of additional systemic and hormonal therapies have shown promise in select patients with a particularly high risk of recurrence.^2–4^ The current challenge remains risk stratifying PCa patients to identify patients who would most benefit from more aggressive treatment.

The National Comprehensive Cancer Network (NCCN) risk grouping system is among the most widely used methods to risk stratify PCa patients.^5^ Often simplified into a three-group system (low-, intermediate-, and high-risk) based on prostate specific antigen (PSA), Gleason Grade Sum (GS), and clinical TNM staging, the NCCN risk grouping provides some basic estimation of disease aggressiveness, but has shown suboptimal prognostic and discriminatory ability.^6–8^ Tissue-based genomic and pathologic classifiers using biopsy samples have shown promise at providing an additional layer of risk stratification but have had limited adoption in part because of costs, laboratory requirements, and processing time.^9–11^ Moreover, in the pre-treatment setting, both the NCCN risk grouping and tissue based classifiers limit analysis to information obtained from biopsy samples representing only a small percentage of the prostate and may be influenced by sampling error.^12–16^

Multiparametric MRI has increasingly been employed in the diagnosis and treatment of PCa and potentially allows for comprehensive analysis of the entire prostate as well as potential cancer spread to surrounding pelvic structures.^17–19^ Previous studies which have shown success in assessing PCa risk from prostate MRI have focused primarily on radiologist scoring of prostate MRIs. Such methods require considerable expertise and are still subject to inter-observer variability.^20,21^ More objective machine learning approaches using textual radiomic features of suspicious lesions within the prostate have also shown prognostic ability; however, they are limited by subjective tumor delineation and are of no value in the estimated 15-18% of PCa patients without discernable lesions on MRI.^22–25^ Lastly, analysis of only individual prostate lesions fails to account for the proximity of disease to the prostate capsule or the presence of disease in the surrounding seminal vesicles.^26–37^

One possible solution to help leverage MRI for PCa risk stratification is through deep learning analyses of the entire prostate gland and surrounding seminal vesicles. Deep learning using convolutional neural networks has demonstrated the ability to derive prognostic imaging biomarkers from diagnostic imaging for a variety of malignancies including head and neck cancer, breast cancer, and lung cancer. ^38–42^

In this study, we use deep learning to develop and externally validate two MRI imaging biomarkers for extra-prostatic extension (EPE) and seminal vesicle invasion (SVI) which can aid in the risk stratification of localized PCa. We demonstrate that both biomarkers can independently assess risk of biochemical recurrence in PCa patients treated with local therapy.

Lastly, we demonstrate how the integration of these biomarkers into the existing clinicopathologic risk groupings improve our ability to identify high risk PCa phenotypes.

## Results

### Study Population

We developed two deep learning models to identify EPE and SVI on prostate MRI and validated the models in three held-out datasets representing different practice settings, imaging protocols, and MRI manufacturers. For training and internal validation, a total of 820 patients with biopsy proven PCa treated at Yale were included in the analysis. A total of 612 (75%) and 208 (25%) patients were randomly assigned to the training and internal test sets, respectively. Table 1 provides a summary of the baseline clinical and disease characteristics of all patients in the primary cohort. The average age at diagnosis was 66.1 years (interquartile range (IQR): 60-72), the average time between acquisition of MRI and primary treatment was 137.5 days (IQR: 83.5-173), and the average follow-up was 3.14 years (IQR: 1.57 – 4.42). A balanced representation of treatment modalities was observed; a total of 397 patients (48.4%) received surgery and 423 patients (51.6%) received radiation.

**Table 1.** Baseline Demographics and Disease Characteristics of Training and Validation Cohorts.

|  | Radical Prostatectomy |  |  | Radiation Therapy |  |  |
| --- | --- | --- | --- | --- | --- | --- |
|  | Training | Validation | P | Training | Validation | P |
| **N** | 290 | 107 |  | 322 | 101 |  |
| **Age, mean (SD)** | 62.2 (6.9) | 61.9 (7.6) | 0.50 | 70.3 (7.7) | 68.6 (7.4) | 0.25 |
| **PSA (ng/mL), mean (SD)** | 9.8 (12.1) | 9.9 (10.7) | 0.47 | 17.4 (43.7) | 16.8 (36.5) | 0.27 |
| **MRI acquisition to tx time, days, mean (SD)** | 110.8 (82.5) | 117 (77.3) | 0.24 | 162.4 (94.1) | 157.6 (97.4) | 0.49 |
| **Race, n (%)** |  |  | 0.91 |  |  | 0.94 |
| White | 236 (81.4) | 88 (82.2) |  | 253 (78.6) | 79 (78.2) |  |
| Black or African-American | 34 (11.7) | 11 (10.3) |  | 51 (15.8) | 17 (16.8) |  |
| Other | 20 (6.9) | 8 (7.5) |  | 18 (5.6) | 5 (5.0) |  |
| **Gleason Grade Sum, n (%)** |  |  | 0.77 |  |  | 0.33 |
| 6 | 22 (7.6) | 6 (5.6) |  | 5 (1.6) | 2 (2.0) |  |
| 7 | 185 (63.8) | 71 (66.4) |  | 192 (59.6) | 68 (67.3) |  |
| 8+ | 83 (28.6) | 30 (28.0) |  | 125 (38.8) | 31 (30.7) |  |
| **Percent Positive Biopsy Cores, n (%)** |  |  | 0.48 |  |  | 0.55 |
| < 34% | 100 (34.5) | 30 (28.0) |  | 89 (27.6) | 27 (26.7) |  |
| 34 - 50% | 72 (24.8) | 29 (27.1) |  | 95 (29.5) | 25 (24.8) |  |
| ≥ 50% | 118 (40.7) | 48 (44.9) |  | 138 (42.9) | 49 (48.5) |  |
| **Clinical Stage, n (%)** |  |  | 0.62 |  |  | 0.59 |
| cT1 | 217 (74.8) | 81 (75.7) |  | 227 (70.5) | 66 (65.3) |  |
| cT2 | 55 (19.0) | 22 (20.6) |  | 49 (15.2) | 19 (18.8) |  |
| cT3 | 18 (6.2) | 4 (3.7) |  | 46 (14.3) | 16 (15.8) |  |
| **NCCN Risk Group, n (%)** |  |  | 0.85 |  |  | 0.77 |
| Low | 11 (3.8) | 3 (2.8) |  | 2 (0.6) | 1 (1.0) |  |
| Intermediate-favorable | 61 (21.0) | 21 (19.6) |  | 51 (15.8) | 16 (15.8) |  |
| Intermediate-unfavorable | 117 (40.3) | 48 (44.9) |  | 121 (37.6) | 43 (42.6) |  |
| High/Very-High | 101 (34.8) | 35 (32.7) |  | 148 (46.0) | 41 (40.6) |  |
| **CAPRA Score, n (%)** |  |  | 0.80 |  |  | 0.18 |
| 0-2 | 38 (13.1) | 10 (9.3) |  | 15 (4.7) | 5 (5.0) |  |
| 3-5 | 176 (60.7) | 66 (61.7) |  | 158 (49.1) | 58 (57.4) |  |
| 6-8 | 72 (24.8) | 28 (26.2) |  | 130 (40.4) | 29 (28.7) |  |
| 9-10 | 7 (2.4) | 3 (2.8) |  | 19 (5.9) | 9 (8.9) |  |
| **Pathological Stage, n (%)** |  |  | 0.99 |  |  |  |
| pT2 | 134 (46.2) | 51 (47.7) |  | .. | .. |  |
| pT3a | 106 (36.6) | 40 (37.4) |  | .. | .. |  |
| pT3b | 40 (13.8) | 15 (14.0) |  | .. | .. |  |
| NA | 10 (3.4) | 0 (0.0) |  | 322 (100.0) | 101 (100.0) |  |
| **Surgical Margins, n (%)** |  |  | 0.25 |  |  |  |
| Positive | 117 (40.3) | 51 (47.7) |  | .. | .. |  |
| Negative | 165 (56.9) | 54 (50.5) |  | .. | .. |  |
| NA | 8 (2.8) | 2 (1.9) |  | 322 (100.0) | 101 (100.0) |  |
| **Pathological EPE, n (%)** |  |  | 0.90 |  |  |  |
| Positive | 149 (51.4) | 54 (50.5) |  | .. | .. |  |
| Negative | 134 (46.2) | 51 (47.7) |  | .. | .. |  |
| NA | 7 (2.4) | 2 (1.9) |  | 322 (100.0) | 101 (100.0) |  |
| **Pathological SVI, n (%)** |  |  | 0.92 |  |  |  |
| Positive | 44 (15.2) | 15 (14.0) |  | .. | .. |  |
| Negative | 242 (83.4) | 90 (84.1) |  | .. | .. |  |
| NA | 4 (1.4) | 2 (1.9) |  | 322 (100.0) | 101 (100.0) |  |
| **Radiologic EPE, n (%)** |  |  | 0.92 |  |  | 0.86 |
| Positive | 29 (10.0) | 12 (11.2) |  | 51 (15.8) | 16 (15.8) |  |
| Negative | 187 (64.5) | 67 (62.6) |  | 181 (56.2) | 54 (53.5) |  |
| Probable or Possible | 74 (25.5) | 28 (26.2) |  | 90 (28.0) | 31 (30.7) |  |
| **Radiologic SVI, n (%)** |  |  | 0.10 |  |  | 0.06 |
| Positive | 16 (5.5) | 4 (3.7) |  | 32 (9.9) | 4 (4.0) |  |
| Negative | 269 (92.8) | 97 (90.7) |  | 279 (86.6) | 90 (89.1) |  |
| Probable or Possible | 5 (1.7) | 6 (5.6) |  | 11 (3.4) | 7 (6.9) |  |

BCR: Biochemical Recurrence, PSA: Prostate Specific Antigen, EPE: Extraprostatic Extension, SVI: Seminal Vesicle Invasion, tx: Treatment

There were no significant differences observed between the training and internal test sets across all clinicopathologic variables; this was consistent when stratifying by treatment modality (Table 1). On the other hand, there were significant differences in the clinicopathologic composition of patients between surgery and radiation subsets (Figure 2c). Consistent with national trends, patients who received radiation were older at diagnosis (p < 0.001, MWU-test), had higher diagnostic PSA levels (p < 0.001, MWU-test), had higher biopsy GSs (p < 0.001, χ^2^ test), more advanced clinical T-Stage (p < 0.001, χ^2^ test), higher Cancer of the Prostate Risk Assessment (UCSF-CAPRA) Scores (p < 0.001, χ^2^ test), and higher NCCN risk categories (p < 0.001, χ^2^ test). ^6,43^

**Fig. 1.**
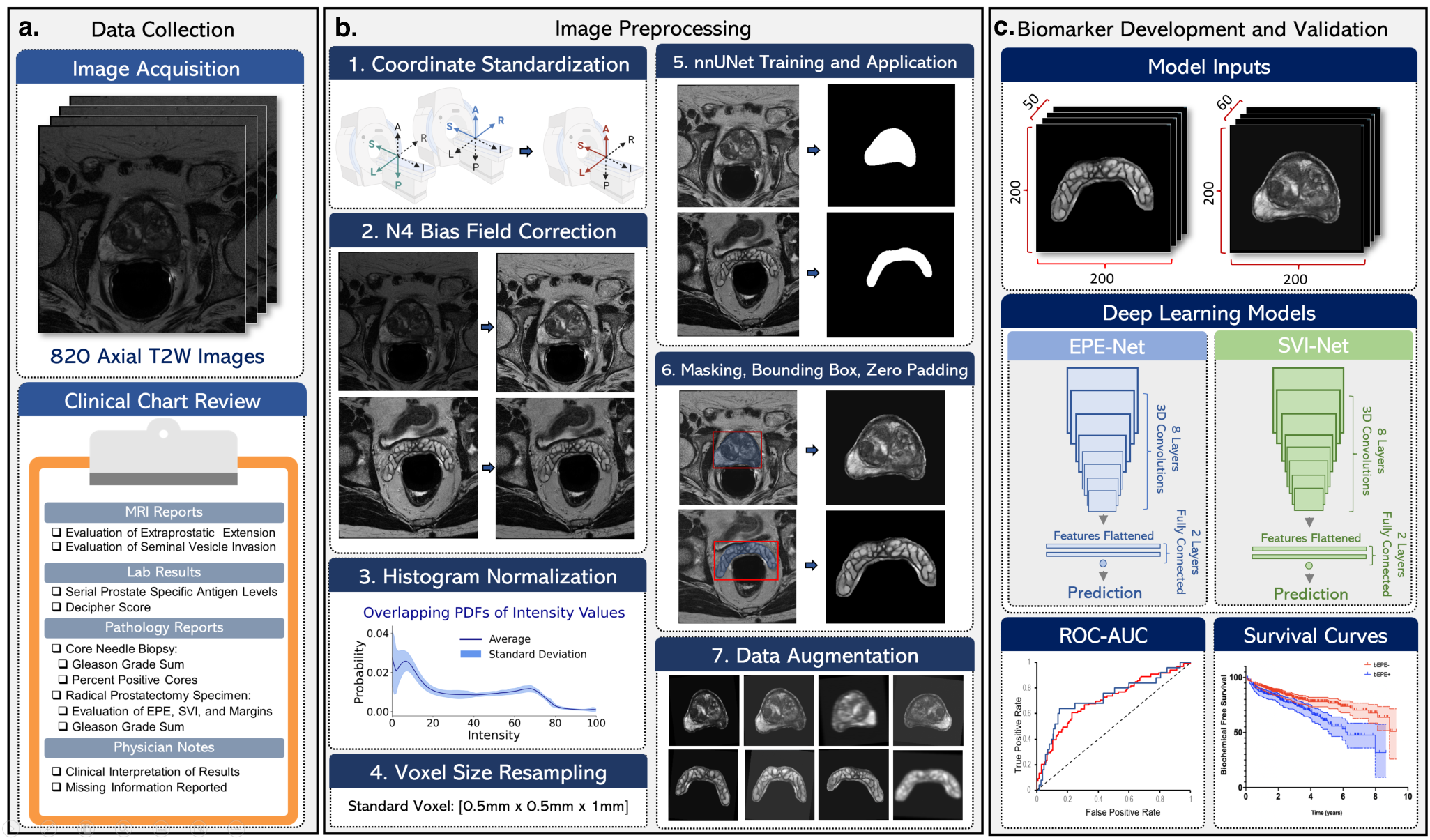
Overview of overall project workflow and image preprocessing pipeline. **a** The primary cohort for this study included n=820 biopsy-proven prostate cancer patients who were treated with either surgery or radiation at our institution between 2010 and 2021. Axial T2W MRI images of the pelvis/prostate for each patient were deidentified and transferred to a secure research PACS for processing. Clinical data was collected for each patient by way of retrospective chart review, with a focus on obtaining an MRI report with evaluation of Extra-prostatic Extension (EPE) and Seminal Vesicle Invasion (SVI) for each image, serial Prostate-Specific Antigen (PSA) labs for pre- and post-treatment evaluations, pathology reports of core needle biopsy specimens and surgical specimens with evaluation of core positivity, Gleason Grade, EPE, SVI, and Surgical Margin Status (SMS) and physician notes to obtain staging, interpretation of results, and missing information not found directly in the chart. **b** Pipeline for image preprocessing included standardization of all images to the LAS axis orientation, N4 bias field correction to correct for B1-field variations within an image and inhomogeneity between different images, intensity normalization performed by the method proposed by Nyul et al. (1999) where a set a learned histogram features across a subset of images is matched across all images, and resampling to a standard voxel size of 0.5mm×0.5mm×1mm. nnU-Net is trained on a random subset of 60-100 manually-contoured prostate and seminal vesicles, and the trained algorithm is used to run segmentation inferences across all other images. Images are then masked with prostate and seminal vesicle segmentations, and zero padding is applied to generate two separate 3D volumes for each patient: a seminal vesicle volume with a size of 100mm×100mm×50mm and a prostate volume with a size of 100mm×100mm×60mm. Prior to training deep learning models, extensive data augmentation is performed. **c** Two deep learning models for EPE and SVI are developed to generate two separate imaging biomarkers. The architecture for each network is roughly equivalent, including eight 3D convolutional neural network layers for feature extraction and a two layer fully-connected network to make a prediction from 2,304 flattened features. Biomarkers are generated by generating Receiver-Operating Curves (ROC) and measuring Area Under the Curve (AUC) across three different test sets, and Kaplan-Meier survival analysis is performed for each biomarker with respect to predicting biochemical recurrence (BCR).

**Fig. 2.**
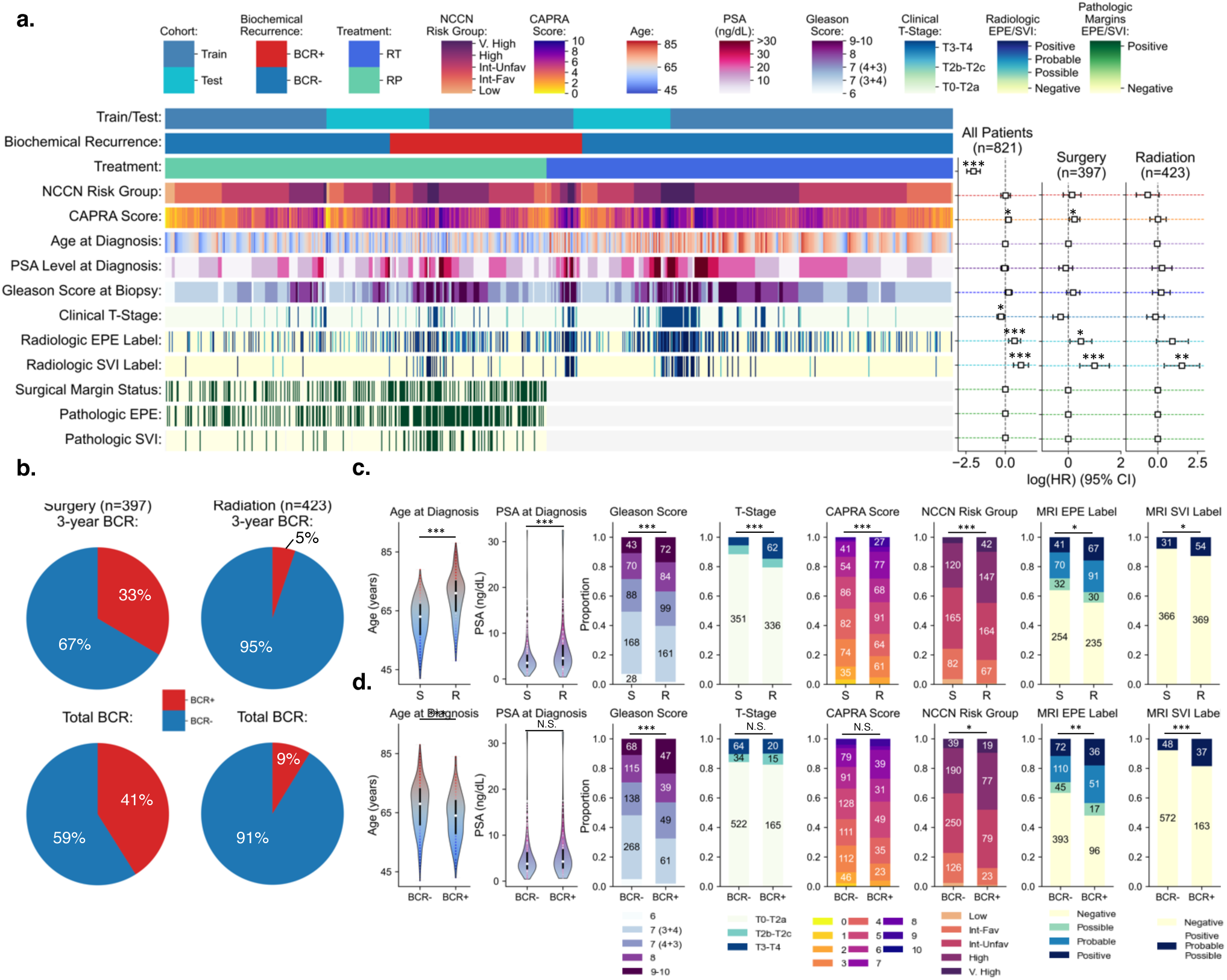
Characteristics and risk group categories of primary training and test cohorts. **a** Heatmap listing the NCCN and CAPRA risk group classifications and the clinical, pathological, and radiologic characteristics for n=820 patients, split into n=612 and n=209 in training and testing cohorts, respectively. Deep learning models were trained using radiologic EPE and SVI labels determined based on a four-point probabilistic translation of readings made by expert abdominal radiologists at our institution, depicted in rows 10 and 11 of the heatmap with range of values including negative, possible, probable, and positive. On the right are adjusted log hazard ratios (HRs) for biochemical recurrence (BCR) based on multivariate Cox Proportional Hazard (CoxPH) models across all patients and stratified by treatment modality; the rows of each model correspond to the adjacent row in the heatmap to the left. For all CoxPH models, *** denotes p < 0.005, ** denotes p < 0.01, and * denotes p < 0.05 based on a likelihood-ratio test. The ground-truth radiologic labels were significantly associated with BCR across all patients (n=820), with HRs of 0.58 (95% CI: 0.20-0.96; p < 0.005) for EPE and 0.99 (95% CI: 0.51-1.47; p < 0.005) for SVI. This association is maintained when stratifying patients based on treatment modality; amongst surgery patients (n=397), the HRs are 0.48 (95% CI: 0.06-0.90; p = 0.02) and 1.00 (95% CI: 0.44-1.56; p < 0.005), and for radiation patients (n=423), the HRs are 0.92 (95% CI: -0.09-1.92; p = 0.07) and 1.52 (95% CI: 0.40-2.63; p = 0.01) for radiologic EPE and SVI, respectively. **b** Piecharts showing the 3-year and overall rates of BCR across surgery (left) and radiation (right) patients. BCR was defined as a PSA > 0.1 ng/mL after treatment, for patients treated with surgery, and a PSA rise above nadir ³ 2 ng/mL, for patients treated with radiation therapy. The 3-year rates of BCR were 33% and 5%, and the overall rates of BCR were 41% and 9% for surgery and radiation patients, respectively. **c** Differentiating clinical characteristics of patients treated with surgery and radiation depicted as violin plots for continuous variables and stacked bar plots for categorical variables. Across all x-axes, ‘S’ is an abbreviation for surgery patients and ‘R’ is an abbreviation for radiation patients. For violin plots, the interior black bar represents the IQR (25-75%), and the median is represented as a white dot. *** denotes p < 0.001, ** denotes p < 0.01, and * denotes p < 0.05 based on two-sided Mann-Whitney U-tests for continuous variables and chi-squared tests of proportions for categorical variables. Radiation patients have higher age at diagnosis (p = 7.31×10^-42^), PSA levels at diagnosis (p = 9.40×10^-9^), Gleason Scores (p = 2.92×10^-4^), Clinical T-Stage (p = 3.36×10^-5^), CAPRA Scores (p = 1.36×10^-5^), NCCN Risk Groups (p = 1.82×10^-3^), and greater proportions of non-negative radiologic EPE labels (p = 0.029) and SVI labels (p = 0.027). **d** Differentiating clinical characteristics of patients presenting with and without BCR. Patients who developed BCR have lower age at diagnosis (p = 8.55×10^-8^) but higher Gleason Scores (p = 2.30×10-5), NCCN Risk Groups (p = 0.011), and have greater proportions of non-negative radiologic EPE labels (p = 1.35×10^-3^) and SVI labels (p = 2.59×10^-5^).

In addition to the internal test set, two external real-world test sets were used to validate the EPE and SVI classification models. The first, a set of patients (n=65) treated in non-academic community settings (Com. Hosp.), and the second, a publicly available set of patients (n=41) who underwent low resolution MRI as a part of the NCI-ISBI Prostate Diagnosis Challenge (Prostate-Dx).^44^

Ground-truth labels for both EPE and SVI for all samples were found using a combination of pathology and radiology reports. All ground truth labels were further validated by two board-certified radiologists and one board-certified radiation oncologist (See Methods). For development of an imaging biomarker for EPE, there was an adequate representation of patients (n=331, 40.4%) with EPE across training and test sets. In contrast, SVI was a rarer phenotype (n= 85, 10.4%) (Table 1).

### EPE and SVI Association with BCR

Biochemical recurrence (BCR) was a secondary outcome of interest to assess the prognostic utility of our models. Overall, 24.4% (n=200 patients) developed BCR representing 41.0% of patients treated with surgery (n=163) and 8.2% of patients treated with radiation (n=37) (Supplementary Table 1). BCR was associated with lower age at diagnosis (p < 0.001, MWU-test), higher biopsy GSs (p < 0.001, χ^2^ test), and higher NCCN risk categories (p = 0.003, χ^2^ test), while differences in diagnostic PSA levels (p = 0.079, MWU-test), clinical T-Stage (p = 0.58, χ^2^ test) and UCSF-CAPRA scores (p = 0.076, χ^2^ test) were not significant (Figure 2d). The presence of EPE (p < 0.001, χ^2^ test) and SVI (p < 0.001, χ^2^ test) ground truth labels was associated with increased rates of BCR suggesting prognostic significance. (Figure 2d).

Furthermore, when adjusting for clinical covariates within a Cox Proportional Hazard (CoxPH) model, EPE and SVI ground truth labels remained associated with BCR. (Figure 2a). The CoxPH model resulted in a Harrell’s C-index of 0.80 and the EPE and SVI labels were the most significant features associated with BCR, with adjusted Hazard Ratios (aHRs) of 1.76 (95% confidence interval (95% CI): 1.20 – 2.58) (p < 0.005, likelihood-ratio test) and 2.72 (95% CI: 1.69 – 4.40) (p < 0.005, likelihood ratio test), respectively. The associations remained significant when stratifying these models by treatment type. For surgery patients, the C-index was 0.71; the aHR for EPE was 1.59 (95% CI: 1.06 – 2.41) (p = 0.026, likelihood-ratio test) and was 2.91 (95% CI: 1.67 – 5.07) (p < 0.005, likelihood-ratio test) for SVI. For radiation patients the C-index was 0.74; the aHR for EPE was 3.11 (95% CI: 1.11 – 8.70) (p = 0.031, likelihood-ratio test) and was 3.45 (95% CI: 1.14 – 10.52) (p = 0.030, likelihood-ratio test) for SVI.

### Identification of EPE using Deep Learning

The trained EPE model demonstrated good discriminatory ability in identifying EPE across the three held-out test sets. On the Yale test set, the EPE classifier model achieved an area under the receiver operating characteristic curve (AUC) of 0.719 (95% CI: 0.601 – 0.817). The EPE model validated on the two external datasets, achieving AUCs of 0.743 (95% CI: 0.619 – 0.858) and 0.611 (95% CI: 0.435 – 0.786) on the Com. Hosp. and Prostate-Dx test sets, respectively (Figure 3b). During subset analysis among surgical patients with final pathology, the EPE model maintained similar performance achieving an AUC of 0.666 (95% CI: 0.610-0.719). Similarly, the EPE model maintained consistent performance across different clinical subgroups (Supplemental Table 4, Supplemental Fig 5).

**Fig. 3.**
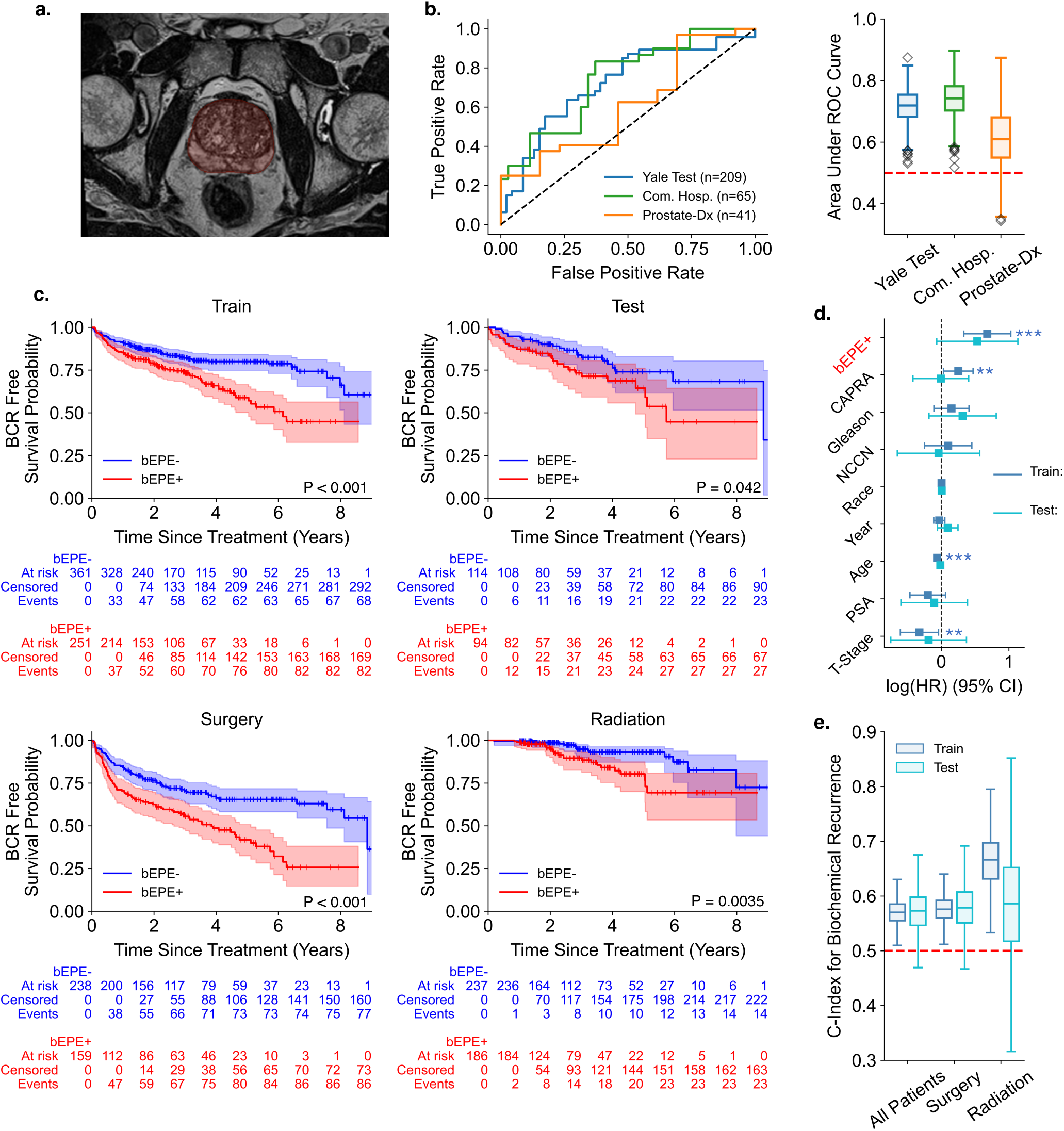
Validation of a bioimaging marker for EPE (bEPE) across cohorts and prediction of biochemical recurrence. **a** T2-Weighted Axial MRI image of prostate with overlaying nnU-Net segmentation of the whole prostate gland highlighted in red. **b** Receiver-Operating Characteristic (ROC) curves of the EPE model across three different test sets (left) and box-and-whisker plots of 95% CI of Area Under the ROC Curve (AUC) constructed with 1,000 bootstrapped resamples of each test set. **c** Kaplan-Meier survival analysis, with P values obtained from the log-rank test, comparing patients with the bioimaging marker for EPE, bEPE+ in red, to those without, bEPE-in blue, as determined from the output of the EPE model, demonstrates significant separation of curves for all patients in the train cohort (p = 2.26×10^-5^), test cohort (p = 0.042), and stratified by patients treated with surgery (p = 6.86×10^-6^) and radiation (p = 3.48×10^-3^). **d** HRs for BCR based on multivariate CoxPH models, stratified by training and test cohorts, demonstrate a significant association for patients positive for bEPE marker with HR of 0.68 (95% CI: 0.33-1.03; p < 0.005) in the training cohort. Across patients in the test cohort, the association is less significant with HR of 0.53 (95% CI: -0.07-1.13; p = 0.08). The vertical dashed line represents a null HR, and P values were obtained from the likelihood-ratio test, with ***, **, and * meaning p < 0.005, p < 0.01, and p < 0.05, respectively. **e** Box-and-whisker plots of 95% CI of C-Indexes for predicting BCR based on bEPE status in training and test cohorts and further stratified by treatment with surgery or radiation. 95% CI were constructed with 10,000 bootstrapped resamples of each cohort.

### Association of EPE Biomarker at BCR

Patients who were classified as having EPE by the EPE model (bEPE+) had significantly increased likelihood of BCR, compared to bEPE-patients. (Figure 3c) In the training set, the 5-year biochemical recurrence free survival (BCR-FS) probabilities were 79.9% for bEPE- and 58.9% for bEPE+ patients (p < 0.001, log-rank test). In the Yale test set, the 5-year BCR-FS probabilities were 74.0% for bEPE- and 64.4% for bEPE+ patients (p = 0.042, log-rank test). For surgery patients, the 5-year BCR-FS probabilities were 65.4% for bEPE- and 41.8% for bEPE+ patients (p < 0.001, log-rank test). Finally, for radiation patients, the 5-year BCR-FS probabilities were 93.1% for bEPE- and 80.3% for bEPE+ patients (p = 0.0035, log-rank test).

The CoxPH model of the bEPE biomarker, adjusting for other clinical covariates, resulted in a C-index of 0.71 with bEPE having an aHR of 1.97 (95% CI: 0.92-0.96). The CoxPH model across the test set resulted in a C-index of 0.65, with bEPE+ having an aHR of 1.71 (95% CI 0.94-3.11) (Figure 3d-e).

### Identification of SVI using Deep Learning

The trained SVI model demonstrated similar discriminatory ability in identifying SVI across the three held-out tests. On the Yale test set, the SVI model achieved an AUC of 0.724 (95% CI: 0.594 – 0.847). The SVI model validated on the two external datasets, achieving AUCs of 0.678 (95% CI: 0.492 – 0.848) and 0.775 (95% CI: 0.485 – 0.990) on the Com. Hosp. and Prostate-Dx test sets, respectively (Figure 4b). During subset analysis among surgical patients with final pathology, the SVI model maintained similar performance achieving an AUC of 0.699 (95% CI: 0.615 – 0.777). Similarly, the SVI model maintained consistent performance across different clinical subgroups (Supplemental Table 5, Supplemental Fig 6).

**Fig. 4.**
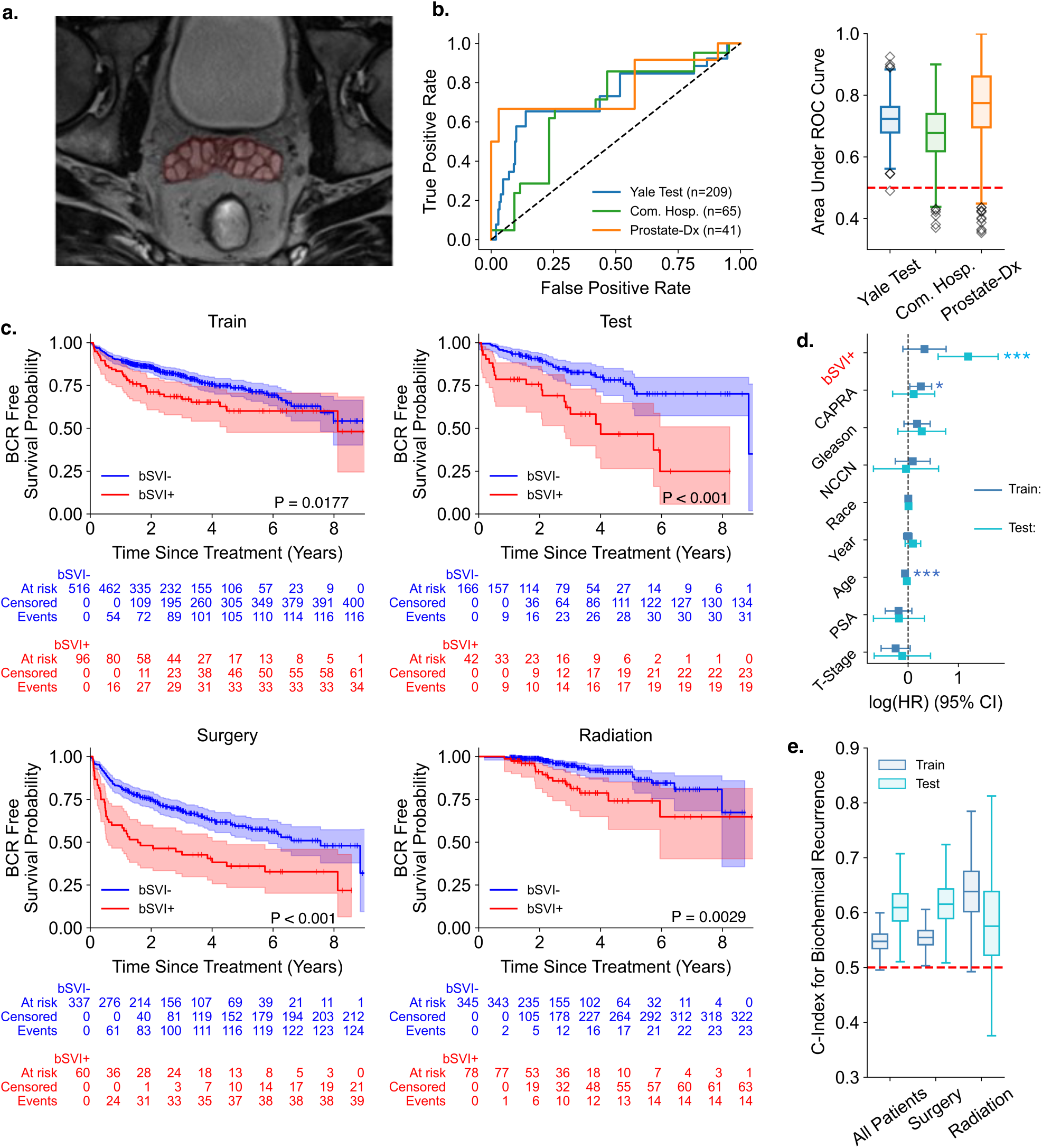
Validation of a bioimaging marker for SVI (bSVI) across cohorts and prediction of biochemical recurrence. **a** T2-Weighted Axial MRI image of seminal vesicles with overlaying nnU-Net segmentation of the whole organ highlighted in red. **b** Receiver-Operating Characteristic (ROC) curves of the SVI model across three different test sets (left) and box-and-whisker plots of 95% CI of Area Under the ROC Curve (AUC) developed with 1,000 bootstrapped resamples of each test set. **c** Kaplan-Meier survival analysis, with P values obtained from the log-rank test, comparing patients with the bioimaging marker for SVI, bSVI+ in red, to those without, bSVI-in blue, as determined from the output of the SVI model, demonstrates significant separation of curves for all patients in the training cohort (p = 0.0177), test cohort (p = 7.84×10^-5^), and stratified by patients treated with surgery (p = 2.14×10^-5^) and radiation (p = 2.94×10^-3^). **d** HRs for BCR based on multivariate CoxPH models, stratified by training and test cohorts, demonstrate a significant association for patients positive for bSVI marker with HR of 1.19 (95% CI: 0.60-1.79; p < 0.005) in the test cohort; however, across patients in the training cohort, the association is less significant with HR of 0.33 (95% CI: -0.10-0.76; p = 0.14). The vertical dashed line represents a null HR, and P values were obtained from the likelihood-ratio test, with ***, **, and * meaning p < 0.005, p < 0.01, and p < 0.05, respectively. **e** Box-and-whisker plots of 95% CI of C-Indexes for predicting BCR based on bSVI status in training and test cohorts and further stratified by treatment with surgery or radiation. 95% CI were constructed with 10,000 bootstrapped resamples of each cohort.

### Association of SVI Biomarker at BCR

Patients who were classified as having SVI by the SVI model (bSVI+) had significantly increased likelihood of BCR, compared to bSVI-patients. (Figure 4c) In the training set, the 5-year BCR-FS probabilities were 73.6% for bSVI- and 60.1% for bSVI+ patients (p = 0.018, log-rank test). In the test set, the 5-year BCR-FS probabilities were 75.8% for bSVI- and 46.6% for bSVI+ patients (p < 0.001, log-rank test). Across all surgery patients, the 5-year BCR-FS probabilities were 59.4% for bSVI- and 36.0% for bSVI+ patients (p < 0.001, log-rank test).

Finally, across all radiation patients, the 5-year BCR-FS probabilities were 91.0% for bSVI- and 74.1% for bSVI+ patients, respectively (p = 0.0029, log-rank test). The CoxPH model of the bSVI biomarker, adjusting for other clinical covariates, resulted in a C-index of 0.71 with bSVI having an aHR of 1.37 (95% CI: 0.89-2.11). The CoxPH model across the test set resulted in a C-index of 0.64, with bSVI+ having an aHR of 3.31 (95% CI 1.82-6.01) (Figure 4d-e).

### Combined EPE/SVI Biomarker and BCR

Combining results from the EPE model and SVI model demonstrated improved prognostic value for BCR. The 5-year BCR-FS probabilities of three groups of patients—those with negative biomarkers (bEPE-& bSVI-), those with only one positive biomarker (bEPE+ or bSVI+), and those with two positive biomarkers (bEPE+ & bSVI+)—were 79.3% (95% CI: 74.7% – 83.8%), 65.6% (95% CI: 51.2% - 80.1%) (p < 0.001, log-rank test), and 60.0% (95% CI: 40.5% - 61.4%) (p < 0.001, log-rank test), respectively (Figure 5a). The C-index for the CoxPH model using the combined imaging biomarker was 0.73, which is an improvement compared to either bSVI or bEPE biomarker alone. Analysis of time-dependent dynamic AUCs for BCR at 1 year, 3 year, and 5 years demonstrates added utility of combining both imaging biomarkers. (Supplementary Table 6) The 5-year C-index for the standalone bSVI and bEPE biomarkers were 0.578 (95% CI: 0.517 – 0.637) and 0.623 (95% CI: 0.563 – 0.683), respectively, whereas the combined imaging biomarker had an improved 5-year C-index of 0.637 (95% CI: 0.583 – 0.690).

**Fig. 5.**
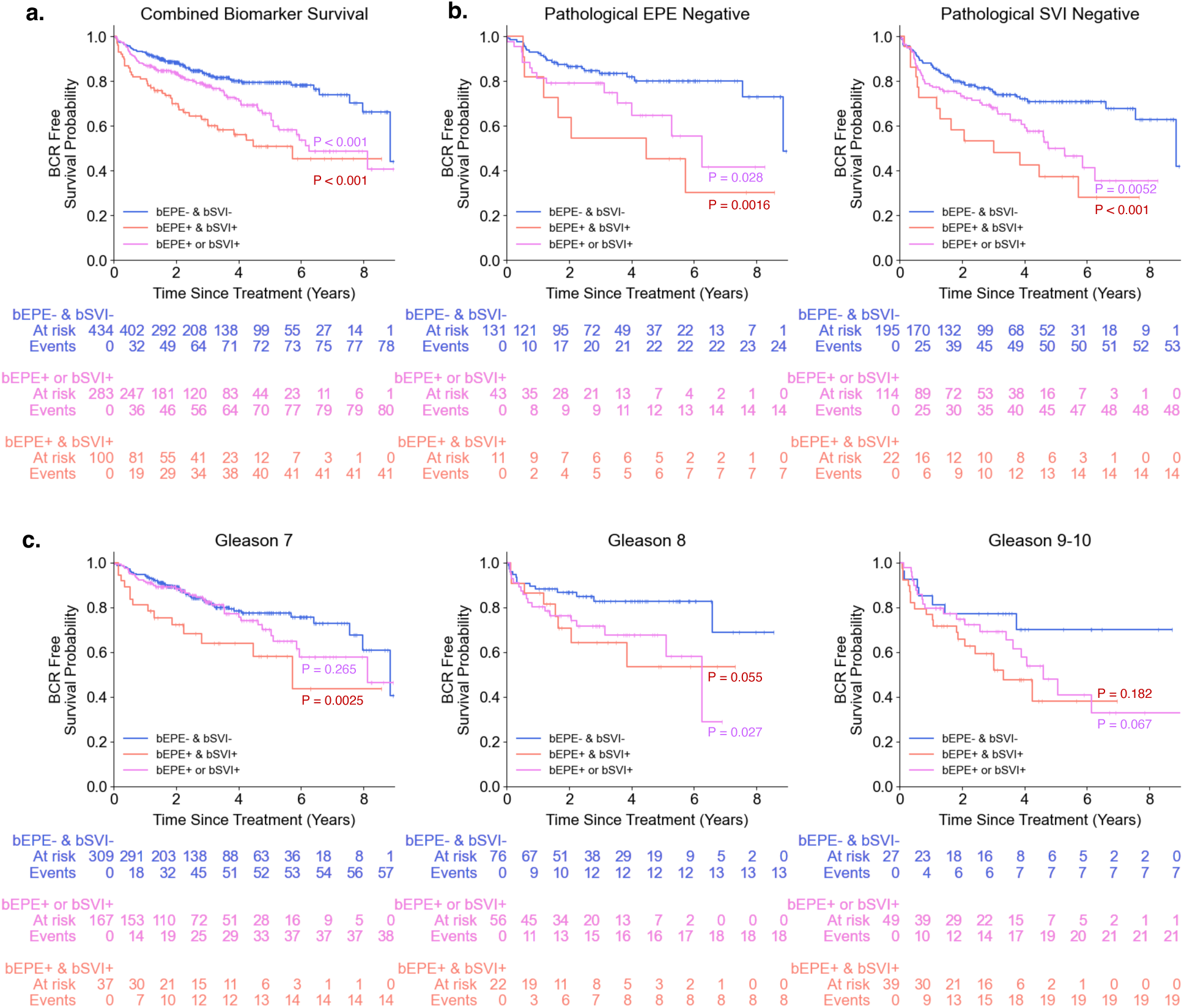
Bioimaging markers for high risk PCa phenotypes can be combined and demonstrate effective risk stratification among pathologically equivalent patients. **a** The bSVI and bEPE can be combined into one system to create three different at-risk groups corresponding to lowest-risk patients who have neither bioimaging marker (bEPE- & bSVI-), intermediate-risk patients who have one or the other but not both (bEPE+ or bSVI+), and the highest risk patients who have both bioimaging markers (bEPE+ & bSVI+). When combined in a Kaplan-Meier survival analysis of biochemical free survival, there is significant separation of curves between each group (p < 0.001). **b** Amongst subsets of patients who received surgery and were discovered to be pathologically negative for EPE (left) and SVI (right), the radiologic-informed biomarkers generated from diagnostic images prior to surgery still demonstrate an ability to distinguish patients at risk for developing BCR, with significant separation of survival curves between low-, intermediate-, and high-risk phenotypes based on our combined biomarker system. **c** Amongst subsets of patients who are homogenous in biopsy Gleason Score, the combined biomarker approach is able to identify patients at greater risk for biochemical recurrence.

When evaluating patients with similar GSs, the combined imaging biomarker was able to successfully identify high/low risk PCa phenotypes associated with BCR. Among Gleason 7 patients, 5-year BCR-FS probabilities were 64.9% with two positive biomarkers, compared to 83.2% in those with two negative biomarkers (p=0.0025, log-rank test). Similarly, among Gleason 8 patients, 5-year BCR-FS probabilities were 63.6% (bEPE+ & bSVI+) vs. 84.2% (p=.055, log-rank test), and among Gleason 9-10 patients, 5-year BCR-FS probabilities were 51.3% (bEPE+ & bSVI+) vs. 74.1% (p=0.182, log-rank test) (Figure 5c).

The combined imaging biomarker approach appears to have utility even among patients without EPE or SVI on final pathology. Among pathologically negative EPE patients (n=185), 5-year BCR-FS probabilities were 45.5% (bEPE+ & bSVI+) vs. 80.2% (95% CI: 72.1% - 88.4%) (p = 0.0016, log-rank test). (Figure 5b, left) In patients without evidence of SVI on final pathology (n=332), 5-year BCR-FS probabilities were 37.3% (bEPE & bSVI+) vs. 70.5% (p < 0.001, log-rank test). (Figure 5b, right) These findings demonstrate the utility of our EPE/SVI biomarkers even among surgical patients for whom EPE and SVI can be pathologically confirmed.

### Interpretability of the EPE and SVI Models

We employed three interpretability experiments to confirm model performance did not stem from heuristics based on spurious data features. (See Methods) First, we used Gradient-weighted Class Activation Mapping (Grad-CAM) to highlight the regions in an image predicting EPE and SVI, respectively.^45^ Grad-CAM heatmaps were compared to radiologist annotations of both EPE and SVI for cases across held-out test sets. For the EPE model, Grad-CAMs tended to localize to the prostatic capsule, demonstrating the model’s aptitude for correctly identifying the location where EPE is found. Grad-CAMs demonstrated concordance with radiologist annotations for both broad capsular EPE as well as more subtle focal EPE (Figure 6b). In contrast, Grad-CAMs for the SVI model highlighted normal seminal vesicle features; activation is seen in regions with high T2 signal intensity, classically associated with normal seminal vesicles. Lower activation was seen in SVI-positive cases due to lower volumes of normal seminal vesicles and loss of T2 signal in the setting of tumor infiltration. (Figure 6e) Representative Grad-CAMs for both correctly and incorrectly classified images from held out test sets are provided in Supplemental Figure 8.

**Fig. 6.**
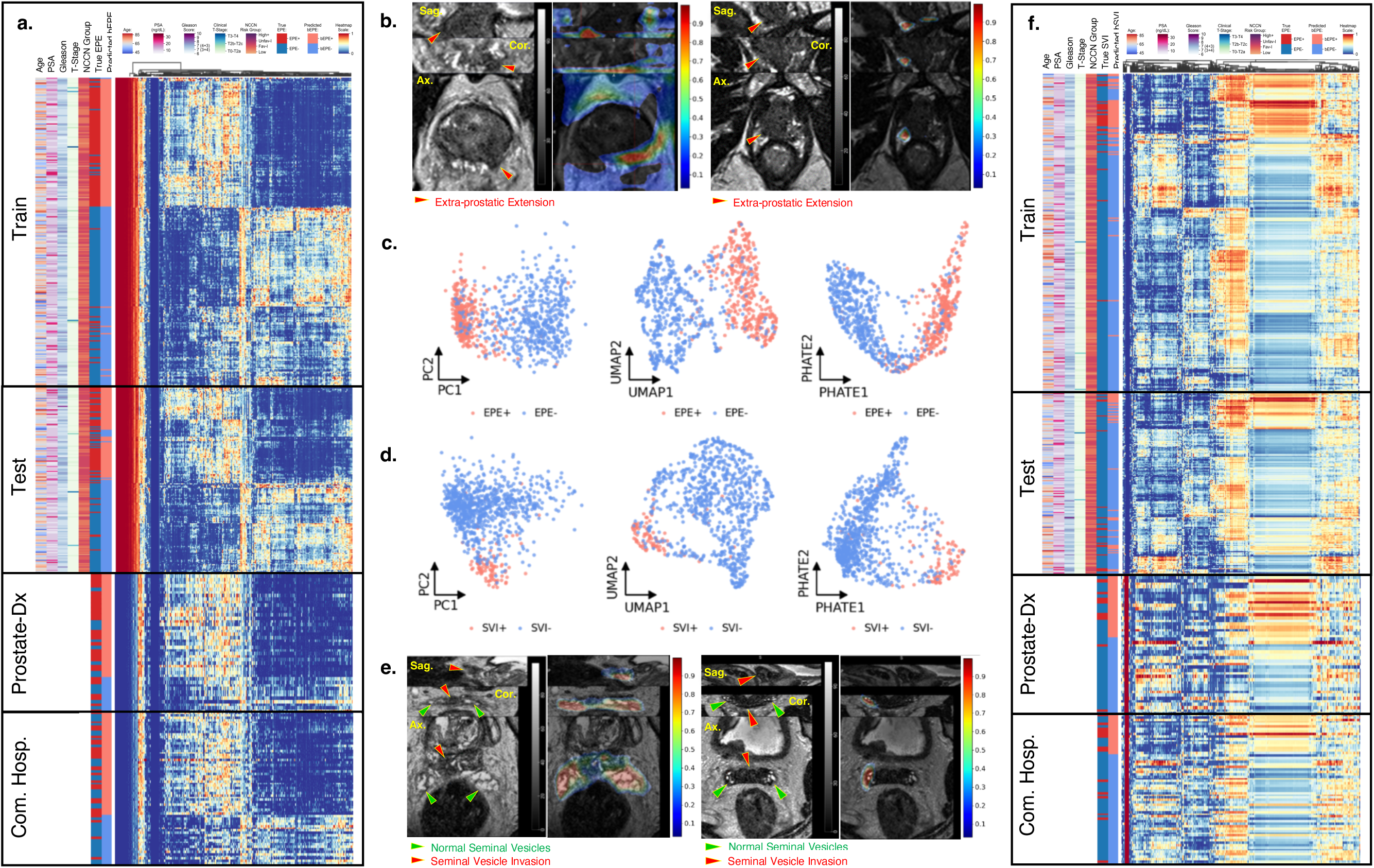
Interpretability of the EPE and SVI models with unsupervised clustering of deeply learned features and Gradient-weighted Class Activation Maps (Grad-CAM). **a,f** Unsupervised hierarchal clustering of normalized deeply learned features (i.e., the output of the flattened features after eight convolutional layers, N=2304) demonstrates two distinct patterns for both EPE (**a**) and SVI models (**f**). For the EPE model, there is a significant association of deep learning expression patterns with true positive EPE across all cohorts (training: p < 0.001, test: p < 0.001, Com. Hosp.: p < 0.001, Prostate-Dx: p < 0.001). For the SVI model, there is also a significant association of deep learning expression patterns with true positive SVI across all cohorts (training: p < 0.001, test: p < 0.001, Com. Hosp.: p < 0.001, Prostate-Dx: p < 0.001). **c, d** Projection of the 2304 flattened features into 2-dimensional space with three different dimensionality reduction algorithms (PCA, UMAP, and PHATE) demonstrate visually distinct separation of communities defined by true EPE status (**c**) and true SVI status (**d**). **b** Grad-CAM visualization of the EPE model demonstrates activation of regions (red on heatmap) with distinct capsular irregularities, suggesting EPE (left), and more subtle regions of capsular contact, suggesting possible EPE. **e** Grad-CAM visualization of the SVI model demonstrates high activation (red on heatmap) in regions with normal seminal vesicle features and low activation (blue on heatmap) in regions with SVI.

Second, unsupervised hierarchical clustering was used to test for associations between model features and evidence of EPE and SVI.^46^ There was a significant association of deep learning feature expression patterns and EPE that persisted when comparing the training and three test sets (Yale train: p < 0.001, Yale test: p < 0.001, Com. Hosp.: p < 0.001, Prostate-Dx: p < 0.001). (Figure 6a) Similarly, for patients with SVI, there was a unique deep learning feature expression pattern which persisted when comparing training and test sets ((Yale train: p < 0.001, Yale test: p < 0.001, Com. Hosp.: p < 0.001, Prostate-Dx: p < 0.001). (Figure 6f) For both EPE and SVI models, deep learning feature expression patterns for EPE were associated with higher NCCN, UCSF-CAPRA scores, Gleason grade group, T-stage, and PSA, suggesting clinical relevance.

Lastly, we employed dimensionality reduction techniques (PCA^47^, UMAP^48^, PHATE^49^) to confirm deep learning features were associated with the presence of EPE and SVI. Across all dimensionality reduction techniques, visually distinct communities defined by EPE and SVI status were identified, suggesting both models were identifying differentiable features related to both EPE and SVI, respectively. (Figure 6c-d)

### Prognostic Utility of Clinicopathologic-Imaging Risk Groups

Given there exists widely-used clinicopathologic grouping to risk stratify patients, we sought to evaluate the added utility of our combined imaging biomarker approach in estimating prognosis by forming clinicopathologic-imaging risk groups. (See methods, Figure 7a)

**Fig. 7.**
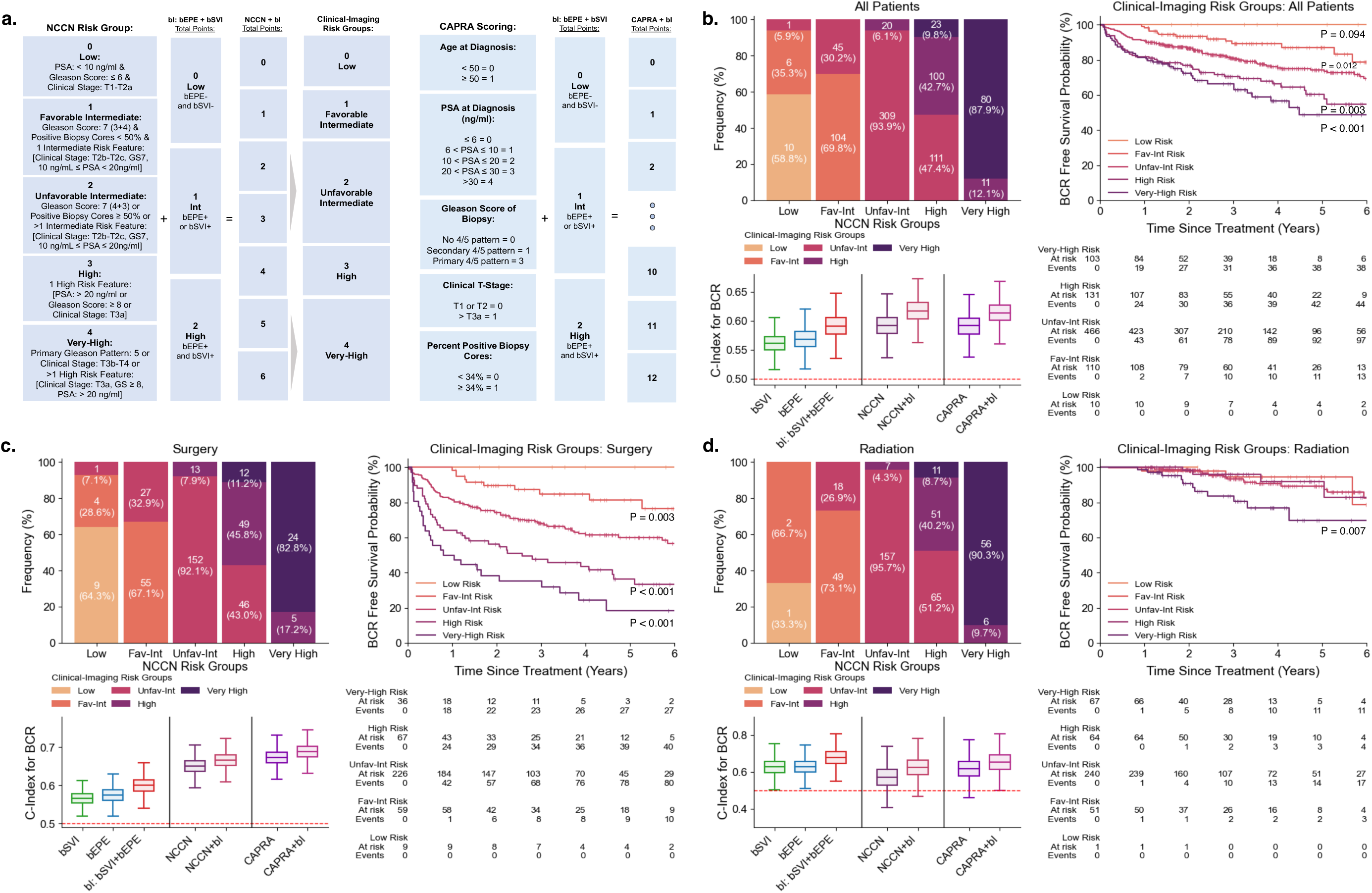
Integrating high risk imaging phenotypes of PCa on MRI into NCCN and CAPRA scoring systems improves risk stratification. **a** NCCN baseline categories are ordered from 0 to 4 points, corresponding to low to very-high risk, and added to the combined biomarker score (bI) ranging from 0-to-2 points. The combined risk system, NCCN+bI, ranges from 0 to 6 points, but subsequent pooling of groups 2 and 3, as well as 5 and 6, create a novel 0-to-4-point reclassification of the original NCCN groups (left). The CAPRA scoring system, ranging from 0 to 10 points, is simply added to the combined biomarker scores to develop a 0-to-12-point stratified system, CAPRA+bI (right). **b** Integrated clinical-imaging risk group system recategorizes 26.5% of all patients (n=217), demonstrates significant heterogeneity within original NCCN risk groups (top left), results in significant separation of curves in Kaplan-Meier (K-M) survival analysis of risk groups (right), and the addition of imaging biomarkers to both NCCN and CAPRA significantly improves C-index for predicting BCR (bottom left). Similar trends are observed for patients who received surgery (**c**); however, for patients who received radiation, (**d**) K-M analysis only demonstrates notable separation for patients classified as very-high risk (right), and although addition of imaging biomarkers does improve C-indexes for both NCCN and CAPRA, the combined biomarker has a better C-index than either NCCN or CAPRA.

Integrating our imaging biomarkers into the NCCN risk groups^6^ resulted in meaningful reclassification of 26.4% (n=217) PCa patients. (Figure 7b-d). 11.6% (n = 95) of the reclassified patients were upgraded to a higher risk group and 14.8% (n = 122) downgraded to a lower risk group. For surgery patients, 27.7% (n = 110) were reclassified, with 14.4% (n = 57) of patients upgraded to higher risk and 13.4% (n = 53) downgraded to lower risk. For radiation patients, 26.8% (n = 109) were reclassified, with 8.9% (n = 38) patients upgraded to higher risk and 16.8% (n = 71) downgraded to lower risk.

Kaplan-Meier (KM) survival analysis for reclassified clinicopathologic-imaging risk groups demonstrated the ability to successfully prognosticate BCR among PCa patients. Across the entire cohort, there was significant separation between KM curves; the 5-year BCR-FS probabilities for low-, favorable-intermediate-, unfavorable-intermediate-, high-, and very-high-risk were 100%, 86.9% (95% CI: 79.6% – 94.2%), 74.3% (95% CI: 62.7% – 85.9%), 60.4% (95% CI: 39.5% – 81.3%), and 48.7% (95% CI: 38.5% - 58.9%), respectively. (Figure 7b)

Specifically, for surgery patients, significant separation of KM curves remained, and the 5-year BCR-FS probabilities were 100%, 81.3% (95% CI: 69.4% – 93.2%), 60.1% (95% CI: 52.7% – 67.4%), 36.4% (95% CI: 7.3% – 65.4%), and 18.2% (95% CI: 0% - 54.8%) (Figure 7c). For radiation patients, very-high-risk patients demonstrated significant separation of KM curves, with 5-year BCR-FS probabilities of 100%, 94.4% (95% CI: 87.5% – 100%), 89.3% (95% CI: 74.1% – 100%), 92.0% (95% CI: 64.4% – 100%), and 69.9% (95% CI: 58.5% -81.3%) (Figure 7d).

Clinicopathologic-imaging risk groups showed improved discriminatory ability compared to clinicopathologic risk groups. (Figure 7b) When studying the NCCN risk groups, the C-index improved from 0.592 (95% CI: 0.552 – 0.631) to 0.617 (95% CI: 0.577 – 0.656) after integrating our combined imaging biomarker. Similarly, when looking at the UCSF-CAPRA risk score, the C-index improved from 0.592 (95% CI: 0.551 – 0.632) to 0.614 (95% CI: 0.574 – 0.653) when combined with our imaging biomarker. This was seen in both patients treated with surgery (Figure 7c) and RT (Figure 7d).

Analysis of time-dependent dynamic AUCs for BCR at 1 year, 3 years, and 5 years across all models demonstrated a similar trend. We observed the addition of imaging biomarkers to clinicopathologic models improved the 5-year AUCs for both NCCN (0.656 vs 0.613) and UCSF-CAPRA (0.651vs 0.621) risk groups. (Supplementary Table 6)

## Discussion

We developed and externally validated algorithms that accurately identified EPE and SVI on MRI in patients with localized PCa. The algorithms demonstrated high discrimination across clinical patient populations and generalized across three different held-out test sets with images acquired across various clinical settings, using different equipment manufacturers and magnet strengths. The models demonstrated prognostic significance and improved current risk stratification schemes commonly used to identify high risk PCa phenotypes. Our approach has the potential to help personalize PCa treatment and better identify patients who may benefit from intensification and de-intensification of treatment regimens.

Previous studies have shown radiomic features from individual lesions within the prostate to be associated with the likelihood of EPE^26–30^ and PCa outcomes^32,33,50,51^. In contrast, our deep learning models require no manual feature engineering and evaluate the entire prostate gland and seminal vesicles. Our approach is advantageous because it 1) obliviates the known inter-observer variation associated with identifying lesions on prostate MRI^20,21^ and 2) can still be used for the approximately 15-18% of PCa patients who are without visible lesions on prostate MRI.^22–25,52^ Also, compared single center studies employing deep learning to identify EPE on MRIs^3^, our models have broad external validity across different institutions, MRI manufacturers, and magnet strengths.

Our study highlights growing evidence suggesting MRI imaging can improve the risk stratification of PCa patients otherwise grouped by clinical characteristics.^53–55^ Both our EPE and SVI biomarkers maintained strong prognostic ability for BCR across different ages, races, Gleason scores, and PSA levels. (Supplemental Table 6). We found our combined EPE/SVI MRI biomarker to have improved prognostic performance compared to both traditional NCCN risk groups and UCSF-CAPRA risk scoring. Previous studies combining radiomic features and clinical variables have shown ability to identify high risk phenotypes.^50,51^ In addition to focusing solely on lesions within the prostate, these studies are of limited clinical utility because they do not easily integrate with established risk groupings from the NCCN. Our models may be more informative for treatment decisions because they reframe patients within traditional, well-known risk groups which are used to guide treatment selection.

We found that our MRI biomarkers for EPE and SVI had prognostic utility even among patients who were not found to have pathologic EPE or SVI after surgery. Previous studies of PCa have similarly found that radiologist identification of EPE and SVI have prognostic relevance independent of pathologic tumor stage.^55^ This also suggests that our model could potentially identify hidden features associated with disease aggressiveness, in addition to anatomic location of cancer spread.

Grad-CAM heat maps consistently demonstrated image regions which were relevant in the identification of EPE and SVI. This interpretability may address some of the implementation challenges that affect radiomic models which are not easily interpretable by clinicians.^56,57^ Specifically, Grad-CAMs created by our models may provide greater context to EPE and SVI predictions and may aid in their acceptance among clinicians in routine practice.^58^

We acknowledge the limitations of our study. First, while our models have shown the ability to prognosticate biochemical recurrence, we did not evaluate other clinically relevant end points, such as overall survival and PCa-specific mortality. Prostate MRI has only been integrated into clinical practice over the last 10 years, so it is difficult to identify a large enough cohort of patients who underwent MRI with long-term follow up. With that said, BCR has been shown to be associated with an increased risk of death among PCa patients in multiple trials.^59,60^ Second, our study did not compare our MRI biomarkers to tissue-based classifiers, which have also been shown to be prognostic in patients with localized PCa.^9,13–15^ Adoption of genomic testing for localized PCa has been variable across the United States.^11^ Genomic testing results were not available for most of the patients in our study, given many were treated prior to the adoption of such tests within treatment guidelines. Lastly, although our models performed well on African American patients, they were underrepresented within our study cohort. Based on national trends, African Americans have worse PCa outcomes and are less likely to receive prostate MRIs compared to White men.^61^ Further studies with larger, more diverse patient populations would be necessary to validate our models’ generalizability across all patient groups.

In summary, we developed and externally validated deep learning algorithms which can accurately identify extra-prostatic extension and seminal vesicle invasion on prostate MRI. Our prognostic MRI biomarkers improve risk stratification of localized PCa patients and can aid in the better personalization of PCa treatments.

## Methods

### Data Collection

This study was approved by the Institutional Review Board (IRB) at Yale and collaborating institutions and was compliant with the Health Insurance Portability and Accountability Act (HIPAA). De-identified data, including MRI images, were used and no protected health information was needed for analysis. Clinical and pathological disease characteristics for all patients were collected by way of retrospective chart review, and informed consent was waived due to the retrospective nature of this study.

### Yale Cohort

The primary cohort of this study was selected from a registry of 5,577 patients with prostate adenocarcinoma who were diagnosed or treated at the Smilow Cancer Hospital at Yale-New Haven Hospital between 2004 and 2021. From this registry, patients were selected with strict inclusion and exclusion criteria. The inclusion criteria for this study were as follows: (1) diagnosed with PCa after 2010, (2) diagnosis confirmed by prostate biopsy, (3) pre-treatment MRI image acquired within a year prior to receiving treatment, (4) received primary treatment at Yale, (5) treated with either primary RP or primary RT, (6) had documented BCR or did not have BCR at final follow up, (7) had at least one year of follow-up PSA levels available, except if BCR was recorded prior to one year, and (8) had all necessary clinical information to determine NCCN risk group and UCSF-CAPRA score. The exclusion criteria for this study were as follows: (1) discovered to have PCa only on autopsy, (2) no pre-treatment MRI available, or (3) unavailable MRI report. A total of 820 PCa patients that met the above eligibility criteria were identified and selected for further study. (Table 1)

For all patients in the primary cohort, the clinical variables collected included age at diagnosis, race, PSA level at diagnosis, highest Gleason Grade at biopsy, number and percentage of positive biopsy cores, clinical T-Stage, date of MRI, primary treatment type, date of primary treatment, and serial post-treatment PSA levels. The surgical margin status, pathological EPE (pEPE), and pathological SVI (pSVI) based on the biopsy of RP specimens were collected only for patients treated with surgery. (Figure 1a).

### Test Cohorts

Three different held-out test sets were used to assess the external performance of our deep learning models in identifying the presence of EPE and SVI across different MRI images and acquisition protocols. The deep learning models were blinded to these images during training. The primary Yale test cohort (n = 208) was generated through a random selection of 25% of the Yale cohort. The second test cohort (n=65) contained a selection of diagnostic MRI images of biopsy proven PCa patients from three non-academic community hospitals (Greenwich Hospital, Griffin Hospital, Westerly Hospital), abbreviated “Com. Hosp.” throughout this paper. The third test cohort (n=41) was a subset of images from the publicly available, PROSTATE-DIAGNOSIS (Prostate-Dx) dataset.^44^ Prostate-Dx includes prostate MRI images from Boston University and Radboud University, Nijmegen Medical Center. We included images from this dataset if (1) a radiology report accompanying the MRI image, with descriptions of both EPE and SVI, was available and (2) the patient had no treatment for their PCa or prostate procedure (e.g., TURP) prior to receiving the MRI.

### Ground-Truth Labels

Two independent ground-truth labels, SVI and EPE, were necessary to train two distinct deep learning models for imaging-based biomarker development. For patients who underwent surgery, ground-truth labels were based on final surgical pathology. For patients who underwent primary RT, ground-truth labels were determined based on pre-treatment MRI reports and confirmed by two additional board-certified radiologists and a board-certified radiation oncologist. For cases in which there was disagreement, consensus was determined by majority vote.

The secondary outcome of interest in this study was BCR, which was defined as a post-treatment PSA > 0.1 ng/mL for patients treated with surgery and a PSA rise above nadir ≥ 2 ng/mL for patients treated with RT.^4,62^ If BCR occurred, the time to BCR was calculated by the difference, in days, between the date BCR occurred and the date of surgery or completion of RT.

### MRI Image Acquisition

Pre-treatment axial T2-weighted MRI images of the prostate and seminal vesicles were chosen for our model because they have previously demonstrated the ability to identify clinically significant PCa.^36,37^ Our training and test cohorts included images a across variety of manufacturers, machine models, and magnet strengths. The Yale cohort included only 3T MRI images across three different machine models (Siemens Verio, Siemens Skyra, Siemens Vida).

The Com. Hosp. cohort included a combination of 3T and 1.5T MRI images across two different manufacturers and three different machine models (Siemens Skyra, GE HD-Signa, Siemens Verio). Lastly, the Prostate-Dx cohort included only 1.5T MRI images across a single machine model (Philips Achieva). For all cohorts, images were obtained in the DICOM format, subsequently standardized to the NIfTI file format, and transferred to a secure computing server for preprocessing and analysis.

### Identification of Prostate Gland and Seminal Vesicles

Prostate glands and seminal vesicles were identified using a combination of automated and manual image segmentations and further validated by at least two board-certified radiologists or radiation oncologists. For the Yale cohort, manual segmentations were created from 100 images in the training dataset. Using the 100 manually-segmented images, an nnU-Net auto-segmentation algorithm was trained to perform initial segmentations for the remaining 770 MRI images in the Yale cohort and subsequently reviewed by a board-certified radiologist and a board-certified radiation oncologist. (Supplemental Fig 2) For patients in the Com. Hosp. and Prostate-Dx cohorts, prostate glands and seminal vesicles were identified by manual segmentations and reviewed by a board-certified radiologist and a board-certified radiation oncologist.

### Image Preprocessing

MRI image preprocessing is described in Figure 1b and has been previously validated for prostate MRI.^63^ To avoid data leakage during preprocessing, preprocessing was performed separately for each of the study cohorts, including the training and test sets in the primary study cohort. Image preprocessing included the following steps: (1) All image coordinate systems were reoriented to the standard “LAS” radiologic convention for coordinate systems, meaning more positive x, y, and z coordinate values in the three-dimensional (3D) axis respectively corresponded to greater left, anterior, and superior positions.^64^ (2) Image corrections were performed for B1-field variations using the N4 bias-field correction algorithm.^65^ (3) Intensity values were normalized with histogram matching based on landmarks learned on a subset of images as proposed by Nyul and Udupa et al (1999).^66^ (4) Images were then resampled to an in-plane resolution of 0.5 mm × 0.5 mm with B-Spline interpolation and to a slice thickness of 1 mm with first order resampling.^67^ To reduce possible artifacts generated from resampling, the resolution and slice thickness selected represented the most common of all images in the primary cohort. (5) Pre-trained nnU-Net models, described above, were used to infer segmentations of prostates and seminal vesicles across all images, and results were validated by a board-certified radiologist and a board-certified radiation oncologist .^68^ (6) Separately, the whole prostate gland and seminal vesicle segmentations were dilated by 10 pixels and smoothed with a kernel density estimator (KDE)^69^. (Supplemental Fig 4) The resulting binary image was used to mask over the respective gland on the original image. A bounding box was then applied to the masked images, and zero padding was performed to generate fully preprocessed images with input image sizes of 200 × 200 × 60 voxels for prostates and 200 × 200 × 50 voxels for seminal vesicles. (7) Finally, extensive data augmentation was performed on preprocessed images for the training set including rotation, elastic deformation, flipping, random anisotropy, random bias field, Gaussian noise, Gaussian blurring, and gamma alterations.^70,71^ Examples of preprocessed images across training in test cohorts are included in Supplementary Fig 2.

### Deep Learning Model Development

The architectures for the EPE and SVI deep learning models have a similar conceptual framework, but were trained with different organs of interest, input volume sizes, and ground-truth labels. Both models were composed of eight 3D convolutional layers, followed by a flattening layer and a single fully-connected layer. (Supplemental Fig 3). The models both resulted in a binary output prediction of either EPE or SVI, calculated with a sigmoid activation function of the final logits.^72,73^ Batch normalization was done after every layer and max pooling after every two convolutional layers. All layers were activated with a Leaky ReLu function.^73,74^ Extensive experimentation with increasing/decreasing the number of layers, changing mask dilation, and employing skip connections were tested to ensure optimal deep learning model architectures. (Supplemental Table 2)

### Hyperparameter Optimization

We used a Binary Cross Entropy with Logits Loss Function (BCE-LL) and an adaptive moment (ADAM) optimizer with an initial learning rate (*lr*) of 1e-2, mini-batch size of 4 3D volumes, and a weight-decay of 1e-3.^75^ We also employed an adaptive *lr* scheduler, which halved the *lr* if the training loss failed to decrease over the previous five epochs, configured with a sensitivity of 1e-3 in evaluating the loss differential between epochs and a minimum *lr* of 1e-7. As described above, batch-normalization and dropout were used to mitigate overfitting.

### Data augmentation and correcting for class imbalance

Data augmentation was used during training of both EPE and SVI models to improve generalizability. Transformations were applied to input volumes including combinations of the following single transformations: rotation, elastic deformation, flipping, random anisotropy, random bias field, Gaussian noise, Gaussian blurring, and gamma alterations.^70,71^ These augmented images were static between epochs while training the deep learning models. For the EPE model, data augmentation was performed equally across all training EPE- and EPE+ cases. However, for the SVI model, data augmentation was also utilized as a technique to correct for class imbalance; an approximate 1:10 ratio of SVI+ to SVI-labels was present across our training data. During the start of SVI model training, extensive data augmentation was done only on the SVI+ cases to achieve an initial 1:1 ratio in negative to positive cases. We then used a sequential training method in which the number of data augmentation transformations on SVI+ cases was reduced each sequence to decrease the SVI+ to SVI-ratio from 1:1 to 1:3, then from 1:3 to 1:6, and finally from 1:6 to 1:10 through each remaining training sequence. At the start of each of the SVI model training sequences, the prior model weights and optimizer hyperparameters were loaded to continue the training. Performance across SVI training sequences are included in Supplementary Table 3.

### Model Evaluation Metrics

The trained EPE and SVI models were evaluated on three test sets, described above. The specific metrics used to evaluate model performance included accuracy (ACC), sensitivity (SN), specificity (SP), F1-score (F1), and AUC.^76^ 95% CIs for all metrics are calculated using a 1000-fold bootstrap resampling with replacement method over the respective test set or test subset.^77^ An emphasis was placed on our models’ discriminatory ability, so we primarily report the AUCs throughout the main text of this paper; however, the other metrics can be found in Supplementary Tables 2 and 3. Additionally, receiver operating characteristic (ROC) curves across clinical subgroups from our internal Yale test set can be found in Supplementary Figures 5-6 and Supplemental Tables 4-5. For final models, the prognostic value of the output predictive biomarkers, with respect to predicting BCR across the combined internal training and test set, is also assessed with KM survival analysis, CoxPH models, C-indexes, and time-dependent AUCs for BCR as described in the statistical analysis section.^78–81^

### Interpretability Methods

We employed three interpretability experiments to confirm model performance did not stem from heuristics based on spurious data features.

First, the Grad-CAM method was used to generate heatmaps which localized coarse regions over the input volumes that contributed to the predictions made by the EPE and SVI models.^45^ During backward propagation of each of the trained models, gradients were calculated for each output feature map of the last convolutional layer with respect to the final classification, global average pooling of the gradients was used to determine feature map importance weights, and ReLU activation of the weighted sum of feature maps was performed to generate small Grad-CAM images. The images were then resized and resampled to the original input volume size, intensities were normalized between 0 and 1, and the resulting Grad-CAM image was overlayed with the original volume.

Second, unsupervised hierarchical clustering was used to test for associations between model features and evidence of EPE and SVI.^46^ Input for unsupervised hierarchal clustering was a single vector of 2,304 deeply learned features was extracted by flattening the output feature maps generated from the last convolutional layer from each model. Each of the unique 2,304 features was normalized to a scale of 0 to 1 by subtracting the minimum value and dividing by the maximum value across each cohort. Unsupervised hierarchal clustering of these normalized features was performed on the training set, and the learned feature clustering was subsequently applied to each of the test sets to test for associations.

Lastly, three different dimensionality reduction and two-dimensional visualization techniques were performed on the 2,304 extracted features to test the associations between EPE and SVI. The three algorithms—Principal Component Analysis (PCA), Uniform Manifold Approximation and Projection (UMAP), and Potential of Heat-diffusion for Affinity-based Transition Embedding (PHATE)—were used to demonstrate robust unsupervised separation of positive and negative cases for both EPE and SVI.^47–49^

### Clinicopathologic risk groups

NCCN and UCSF-CAPRA were used as two baseline pre-treatment clinicopathologic risk group systems for comparison of novel integrated imaging biomarkers.^6,43^ Five groups, as defined by the NCCN guidelines, were used to classify low-, favorable-intermediate-, unfavorable-intermediate-, high-, and very-high-risk patients. The NCCN very-low risk group was excluded because treated patients were never very-low risk in our cohort. NCCN guidelines defined low risk factors as PSA (ng/mL) < 10, GS ≤ 6, and tumor stage (cT) of 1 or 2a.

Intermediate risk factors were defined as PSA (ng/mL) ≥ 10 and < 20, GS of 7, and cT of either 2b or 2c. High risk factors were defined as PSA (ng/mL) > 20, GS ≥ 8, and cT of 3a. As per NCCN guidelines, the following risk group definitions were followed. Low risk patients had all three low risk factors. Favorable-intermediate risk patients had GS of 7 (3+4), positive biopsy cores < 50%, and at most, one intermediate risk factor. Unfavorable-intermediate risk patients had GS of 7 (4+3), positive biopsy cores > 50%, or greater than one intermediate risk factor.

High risk patients had only one high risk factor, and very-high risk patients had a GS primary pattern of 5, or a cT between 3b and 4, or greater than one high risk factor. For the UCSF-CAPRA scoring, patients were scored from 0 to 10, based on a sum of points with the following criteria: age at diagnosis < 50 and ≥ 50 received 0 and 1 points, respectively; PSAs (ng/mL) ≤ 6, 6 to 10 inclusive (incl.), 10 to 20 incl., 20 to 30 incl., and > 30 received 0, 1, 2, 3, and 4 points, respectively; GSs with no patterns of 4 or 5, secondary pattern of 4 or 5, and primary pattern of 4 or 5 received 0, 1, and 3 points, respectively; cT of 1 or 2 and cT > 3a received 0 and 1 points, respectively; positive biopsy cores < 34% and ≥ 34% received 0 and 1 points, respectively.^43^

### Developing Clinicopathologic-Imaging Risk Groups

A summation and pooling framework previously employed by Spratt et al. (2018) was employed to integrate our MRI imaging-biomarkers into the NCCN risk grouping to form clinicopathologic-imaging risk groups.^15^ First, the five NCCN risk groups from low-to very-high risk were assigned 0 to 4 points, respectively. Next, biomarker predictions from both the EPE and SVI models were added to the NCCN points, resulting in summed points between 0 and 7.

Finally, points of 2 and 3, as well as 5 and 6, were pooled to regenerate a 0-to-4-point scale that resembled the original NCCN risk groups but contained reclassified patients. For the integration with UCSF-CAPRA, the biomarker predictions from the EPE and SVI models were simply added to the UCSF-CAPRA score, ranging from 0 to 10, to derive a new scoring system between 0 and 12. The combined clinicopathologic-imaging risk group systems are compared to their respective baselines with KM survival analysis, CoxPH models, C-indexes for BCR, and time-dependent AUCs for BCR, as described in the statistical analysis section. For the NCCN based clinicopathologic-imaging model, the summed points between 0 and 7, prior to pooling, were used for C-indexes and time-dependent AUCs to provide the best representation of how the addition of our MRI imaging biomarkers affected risk stratification based on the original NCCN risk groups.

### Statistical Analysis

Comparisons along a variety of clinicopathologic characteristics are made between patients with and without BCR, between treatment with radiation and surgery, and between train and test sets in the primary study cohort using student’s t-test for continuous variables and χ^2^ test for categorical variables.

The specific metrics used to evaluate model performance included ACC, SN, SP, F1, and AUC.^76^ 95% CIs for all metrics are calculated using a 1000-fold bootstrap resampling with replacement method over the respective test set or test subset.^77^

KM survival analysis of BCR-FS, censoring patients who had no reported BCR at their last follow-up, was done to evaluate the prognostic value of our standalone biomarkers and for comparing risk groups in the novel NCCN clinicopathologic-imaging system. For all KM survival analyses, the log-rank test was used to measure the statistical significance between groups.^78^ Multivariate CoxPH models—with covariates including other clinical, pathological, and baseline risk group variables—was performed, with primary input covariates of interest being our ground-truth labels, biomarkers, or novel clinicopathologic-imaging risk group systems, to measure aHRs for developing BCR. For all CoxPH models, the C-index is reported and the likelihood-ratio test is used to measure statistical significance of aHRs.^81^ C-indexes were also calculated, a statistic of the rank correlation of model scores and time to development of BCR, as a measure of the overall discriminatory ability for predicting BCR of our standalone biomarkers, combined biomarker, baseline clinicopathologic risk groups, and our novel modified clinicopathologic-imaging risk groups.^79^ For all models for which C-indexes were calculated, 1,000 iterations of the bootstrap method, resampling with replacement, were performed in parallel to develop paired 95% CIs.

A few approaches were used to compare C-indexes between related, nested-models. First, a distribution of differences of paired C-indexes between a novel model and its baseline model was calculated across all bootstrapped resamples, and the absolute percentage of positive C-index differences was reported. Two statistical methods were used to measure statistical significance between models. The Wilcoxon signed-rank test was used on paired differences of C-indexes across all bootstrapped resamples and 100,000 iterations of random permutation testing between a novel model score and baseline model score was performed for direct comparison.

As a more nuanced measure of the discriminatory performance of all models, cumulative-dynamic time-dependent AUCs were estimated at 1, 3, and 5 years with an inverse probability of censored weighting method as described by Blanche et al. (2013) and 95% CIs were computed from the asymptomatic normality of AUC estimators.^80,82^ All statistical analyses were performed in Python v3.7, and all statistical tests were performed using a .05 significance level.

## Data Availability

MRI data for the Prostate-Dx test dataset is publicly available on The Cancer Imaging Archive. MRI data for the Yale and Community Hospital datasets will be available upon reasonable request to the corresponding author and after data use agreement with associated institutions.

## Code Availability

The MRI image preprocessing code and model code is available on our laboratory GitHub (https://github.com/Aneja-Lab-Yale/Aneja-Lab-Public-Prostate_MRI_Biomarker).

## Supporting information

Suppplemental Material

## Data Availability

All data produced in the present study are available upon reasonable request to the authors

