## Supplementary material for "Deep Learning Identified Extra-Prostatic Extension and Seminal Vesicle Invasion as an MRI Biomarker for Prostate Cancer Outcomes": Suppplemental Material

**Supplementary Table 1:** Baseline Demographics and Disease Characteristics of Primary Yale Train and Test Cohort Stratified by Treatment and Recurrence Status ………………………………………………….. **2**

**Supplementary Figure 1:** Training and Validation Curves for nnU-Net Segmentation of Prostates and Seminal Vesicles …………………………………………………………………………………………………. **3**

**Supplementary Figure 2:** Example Images and Probability Distribution Functions for Preprocessing Steps Across Cohorts ……………..…………………………………………………………………………………. **4**

**Supplementary Figure 3:** Deep Learning Architectures for the EPE and SVI Models…………………….. **5**

**Supplementary Figure 4:** Example Images of Prostate Volumes with Expanded Mask Dilation Kernels for EPE Model Experiments ……………………………………………………………………………………. **6**

**Supplementary Table 2:** The EPE Model Performance Metrics Across Experimental Modifications to Architecture ………..……………………………………………………………………………………………. **6**

**Supplementary Table 3:** The SVI Model Performance Metrics Across Training Sequences …..………... **7**

**Supplementary Table 4:** The EPE Model Performance Across Subgroups of Yale Test Cohort …………………………………………………………………………………………………………………. **8**

**Supplementary Figure 5:** ROC Curves and AUC Boxplots for the EPE Model Across Subsets of Patients …………………………………………………………………………………………………………………. **8**

**Supplementary Table 5:** The SVI Model Performance Across Subgroups of Yale Test Cohort ………..………………………………………………………………..……………………………………. **9**

**Supplementary Figure 6:** ROC Curves and AUC Boxplots for the SVI Model Across Subsets of Patients …………………………………………………………………………………………………………………. **9**

**Supplementary Table 6:** Biomarkers, Clinicopathologic, and Clinicopathologic-Imaging Groups' Time-Dependent AUCs for BCR ………………………………………………………………………………. **10**

**Supplementary Table 7:** Performance of Biomarkers, Clinicopathologic, and Clinicopathologic-Imaging Groups in Predicting Adverse Pathologies in Post-Surgical Specimens ……………………………………. **11**

**Supplementary Figure 7:** ROC Curves and AUC Boxplots for Biomarkers, Clinicopathologic and Clinicopathologic-Imaging Risk Groups in Predicting Adverse Pathologies ………………………………… **12**

**Supplementary Figure 8:** Representative Gradient-Weighted Class Activation Maps (Grad-CAMs) for Correctly and Incorrectly Classified Images ………………………………………………………………… **13**

**Supplementary Table 1. Baseline Demographics and Disease Characteristics of Primary Yale Train and Test Cohorts Stratified by Treatment and Recurrence Status**

| | Radical Prostatectomy | | | | | | Radiation Therapy | | | | | |
|---|---|---|---|---|---|---|---|---|---|---|---|---|
| | Train Cohort | | | Test Cohort | | | Train Cohort | | | Test Cohort | | |
| | **BCR +** | **BCR -** | **P** | **BCR +** | **BCR -** | **P** | **BCR +** | **BCR -** | **P** | **BCR +** | **BCR -** | **P** |
| **N** | 122 | 168 | | 41 | 66 | | 28 | 294 | | 9 | 92 | |
| **Age at diagnosis, mean (SD)** | 62.2 (7.2) | 62.2 (6.8) | 0.97 | 63.1 (6.9) | 61.2 (7.9) | 0.7 | 68.0 (9.6) | 70.5 (7.5) | 0.21 | 68.3 (8.9) | 68.6 (7.3) | 0.96 |
| **pre-tx, PSA (ng/mL), mean (SD)** | 12.6 (17.6) | 7.7 (4.3) | 0.003 | 10.1 (7.6) | 9.7 (12.3) | 0.13 | 41.8 (109.6) | 15.1 (30.4) | 0.1 | 11.0 (5.8) | 17.4 (38.2) | 0.74 |
| **MRI acquisition to tx (days), mean (SD)** | 105.5 (95.5) | 114.6 (71.8) | 0.049 | 106.5 (77.5) | 123.4 (77.0) | 0.057 | 126.2 (68.5) | 165.8 (95.6) | 0.06 | 173.1 (72.2) | 156.1 (99.8) | 0.64 |
| **Race, n (%)** | | | 0.15 | | | 0.91 | | | 0.26 | | | 0.66 |
| White | 93 (76.2) | 143 (85.1) | | 34 (82.9) | 54 (81.8) | | 25 (89.3) | 228 (77.6) | | 8 (88.9) | 71 (77.2) | |
| Black or African-American | 18 (14.8) | 16 (9.5) | | 5 (12.2) | 6 (9.1) | | 3 (10.7) | 48 (16.3) | | 1 (11.1) | 16 (17.4) | |
| Other | 11 (9.0) | 9 (5.4) | | 2 (4.9) | 6 (9.1) | | 0 (0.0) | 18 (6.1) | | 0 (0.0) | 5 (5.4) | |
| **Gleason Grade Sum, n (%)** | | | < 0.001 | | | 0.03 | | | 0.88 | | | 0.26 |
| 6 | 2 (0.0) | 20 (11.9) | | 1 (2.4) | 5 (7.6) | | 0 (0.0) | 5 (1.7) | | 1 (11.1) | 1 (1.1) | |
| 7 | 65 (53.3) | 120 (71.4) | | 24 (58.5) | 47 (71.2) | | 16 (57.1) | 176 (59.9) | | 5 (55.6) | 63 (68.5) | |
| 8+ | 55 (45.1) | 28 (16.7) | | 16 (39.0) | 14 (21.2) | | 12 (42.9) | 113 (38.4) | | 3 (33.3) | 28 (30.4) | |
| **Percent Positive Biopsy Cores, n (%)** | | | < 0.001 | | | 0.79 | | | 0.23 | | | 0.61 |
| < 34% | 27 (22.1) | 73 (43.5) | | 10 (24.4) | 20 (30.3) | | 7 (25.0) | 82 (27.9) | | 3 (33.3) | 24 (26.1) | |
| 34 - 50% | 35 (28.7) | 37 (22.0) | | 12 (29.3) | 17 (25.8) | | 5 (17.9) | 90 (30.6) | | 1 (11.1) | 24 (26.1) | |
| ≥ 50% | 60 (49.2) | 58 (34.5) | | 19 (46.3) | 29 (43.9) | | 16 (57.1) | 122 (41.5) | | 5 (55.6) | 44 (47.8) | |
| **Clinical Stage, n (%)** | | | 0.13 | | | 0.22 | | | 0.52 | | | 0.12 |
| cT1 | 89 (73.0) | 128 (76.2) | | 28 (68.3) | 53 (80.3) | | 18 (64.3) | 209 (71.1) | | 4 (44.4) | 62 (67.4) | |
| cT2 | 22 (18.0) | 33 (19.6) | | 11 (26.8) | 11 (16.7) | | 4 (14.3) | 45 (15.3) | | 4 (44.4) | 15 (16.3) | |
| cT3 | 11 (9.0) | 7 (4.2) | | 2 (4.9) | 2 (3.0) | | 6 (21.4) | 40 (13.6) | | 1 (11.1) | 15 (16.3) | |
| **NCCN Risk Group, n(%)** | | | < 0.001 | | | 0.19 | | | 0.78 | | | 0.26 |
| Low | 1 (0.8) | 10 (6.0) | | 0 (0.0) | 3 (4.5) | | 0 (0.0) | 2 (0.7) | | 1 (11.1) | 0 (0.0) | |
| Intermediate-favorable | 12 (9.8) | 49 (29.2) | | 6 (14.6) | 15 (22.7) | | 5 (17.9) | 46 (15.6) | | 0 (0.0) | 16 (17.4) | |
| Intermediate-unfavorable | 47 (38.5) | 70 (41.7) | | 18 (43.9) | 30 (45.5) | | 9 (32.1) | 112 (38.1) | | 5 (55.6) | 38 (41.3) | |
| High or Very-High | 62 (50.8) | 39 (23.2) | | 17 (41.5) | 18 (27.3) | | 14 (50.0) | 134 (45.6) | | 3 (33.3) | 38 (41.3) | |
| **CAPRA Score, n (%)** | | | < 0.001 | | | 0.07 | | | 0.013 | | | 0.63 |
| 0-2 | 5 (4.1) | 33 (19.6) | | 0 (0.0) | 10 (15.2) | | 2 (7.1) | 13 (4.4) | | 1 (11.1) | 4 (4.3) | |
| 3-5 | 66 (54.1) | 110 (65.5) | | 28 (68.3) | 38 (57.6) | | 8 (28.6) | 150 (51.0) | | 5 (55.6) | 53 (57.6) | |
| 6-8 | 47 (38.5) | 25 (14.9) | | 12 (29.3) | 16 (24.2) | | 13 (46.4) | 117 (39.8) | | 3 (33.3) | 26 (28.3) | |
| 9-10 | 7 (5.7) | 0 (0.0) | | 1 (2.4) | 2 (3.0) | | 5 (17.9) | 14 (4.8) | | 0 (0.0) | 9 (9.8) | |
| **Pathological Stage, n (%)** | | | < 0.001 | | | 0.003 | | | | | | |
| pT2 | 33 (27.0) | 101 (60.1) | | 14 (34.1) | 37 (56.1) | | . . | . . | | . . | . . | |
| pT3a | 53 (43.4) | 53 (31.5) | | 15 (36.6) | 25 (37.9) | | . . | . . | | . . | . . | |
| pT3b | 29 (23.8) | 11 (6.5) | | 12 (29.3) | 3 (4.5) | | . . | . . | | . . | . . | |
| NA | 7 (5.7) | 3 (1.8) | | 0 (0.0) | 1 (1.5) | | 28 (100.0) | 294 (100.0) | | 9 (100.0) | 92 (100.0) | |
| **Surgical Margins, n (%)** | | | < 0.001 | | | 0.1 | | | | | | |
| Positive | 65 (53.3) | 52 (31.0) | | 24 (58.5) | 27 (40.9) | | . . | . . | | . . | . . | |
| Negative | 53 (43.4) | 112 (66.7) | | 16 (39.0) | 38 (57.6) | | . . | . . | | . . | . . | |
| NA | 4 (3.3) | 4 (2.4) | | 1 (2.4) | 1 (1.5) | | 28 (100.0) | 294 (100.0) | | 9 (100.0) | 92 (100.0) | |
| **Pathological EPE, n (%)** | | | < 0.001 | | | 0.02 | | | | | | |
| Positive | 86 (70.5) | 63 (37.5) | | 27 (65.9) | 27 (40.9) | | . . | . . | | . . | . . | |
| Negative | 32 (26.2) | 102 (60.7) | | 13 (31.7) | 38 (57.6) | | . . | . . | | . . | . . | |
| NA | 4 (3.3) | 3 (1.8) | | 1 (2.4) | 1 (1.5) | | 28 (100.0) | 294 (100.0) | | 9 (100.0) | 92 (100.0) | |
| **Pathological SVI, n (%)** | | | < 0.001 | | | < 0.001 | | | | | | |
| Positive | 32 (26.2) | 12 (7.1) | | 12 (29.3) | 3 (4.5) | | . . | . . | | . . | . . | |
| Negative | 88 (72.1) | 154 (91.7) | | 28 (68.3) | 62 (93.9) | | . . | . . | | . . | . . | |
| NA | 2 (1.6) | 2 (1.2) | | 1 (2.4) | 1 (1.5) | | 28 (100.0) | 294 (100.0) | | 9 (100.0) | 92 (100.0) | |
| **Radiologic EPE, n (%)** | | | < 0.001 | | | 0.003 | | | 0.02 | | | 0.81 |
| Positive | 15 (12.3) | 14 (8.3) | | 10 (24.4) | 2 (3.0) | | 9 (32.1) | 42 (14.3) | | 2 (22.2) | 14 (15.2) | |
| Negative | 61 (50.0) | 126 (75.0) | | 21 (51.2) | 46 (69.7) | | 10 (35.7) | 171 (58.2) | | 4 (44.4) | 50 (54.3) | |
| Probable or Possible | 46 (37.7) | 28 (16.7) | | 10 (24.4) | 18 (27.3) | | 9 (32.1) | 81 (27.6) | | 3 (33.3) | 28 (30.4) | |
| **Radiologic SVI, n (%)** | | | 0.003 | | | 0.002 | | | 0.009 | | | 0.08 |
| Positive | 13 (10.7) | 3 (1.8) | | 4 (9.8) | 0 (0.0) | | 7 (25.0) | 25 (8.5) | | 1 (11.1) | 3 (3.3) | |
| Negative | 106 (86.9) | 163 (97.0) | | 32 (78.0) | 65 (98.5) | | 19 (67.9) | 260 (88.4) | | 6 (66.7) | 84 (91.3) | |
| Probable or Possible | 3 (2.5) | 2 (1.2) | | 5 (12.2) | 1 (1.5) | | 2 (7.1) | 9 (3.1) | | 2 (22.2) | 5 (5.4) | |

Abbreviations: **BCR:** Biochemical Recurrence, **PSA:** Prostate Specific Antigen, **EPE:** Extraprostatic Extension, **SVI:** Seminal Vesicle Invasion, **tx:** Treatment, **NCCN:** National Comprehensive Cancer Network, **CAPRA:** Cancer of the Prostate Risk Assessment, **MRI:** Magnetic Resonance Imaging, **N:** Total number of subset cases for the stratification indicated above this value, **n:** Number of cases for indicated demographic or clinical variable to left among total N, **P:** p-values comparing distributions of BCR- and BCR+ cases in the indicated demographic or clinical variable determined by Mann Whitney U-test for continuous variables and $\chi 2$-test for categorical variables.

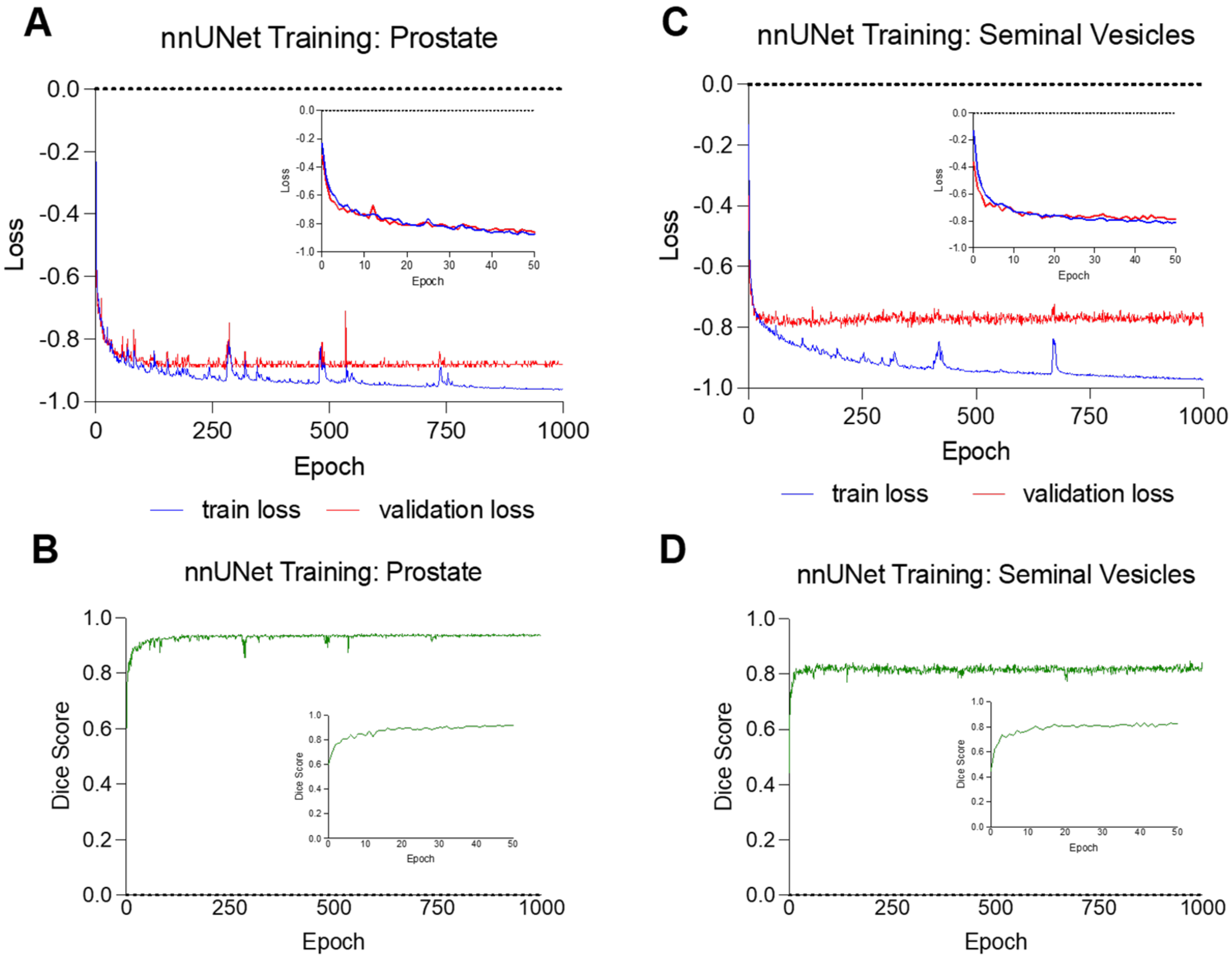


**Supplementary Figure 1 | Training and Validation Curves for nnU-Net Segmentation of Prostates and Seminal Vesicles. a** Loss curve for nnU-Net training on prostate segmentation over 1000 epochs, with interior subpanel showing only the first 50 epochs. **b** Dice score curve on the validation set for nnU-Net training on prostate segmentation task over 1000 epochs, with interior subpanel showing Dice scores across only the first 50 epochs. **c** Loss curve for nnU-Net training on seminal vesicle segmentation over 1000 epochs, with interior subpanel showing loss over only the first 50 epochs. **d** Dice score curve on validation set for nnU-Net training on seminal vesicle segmentation task over 1000 epochs, with interior subpanel showing Dice scores over the first 50 epochs.

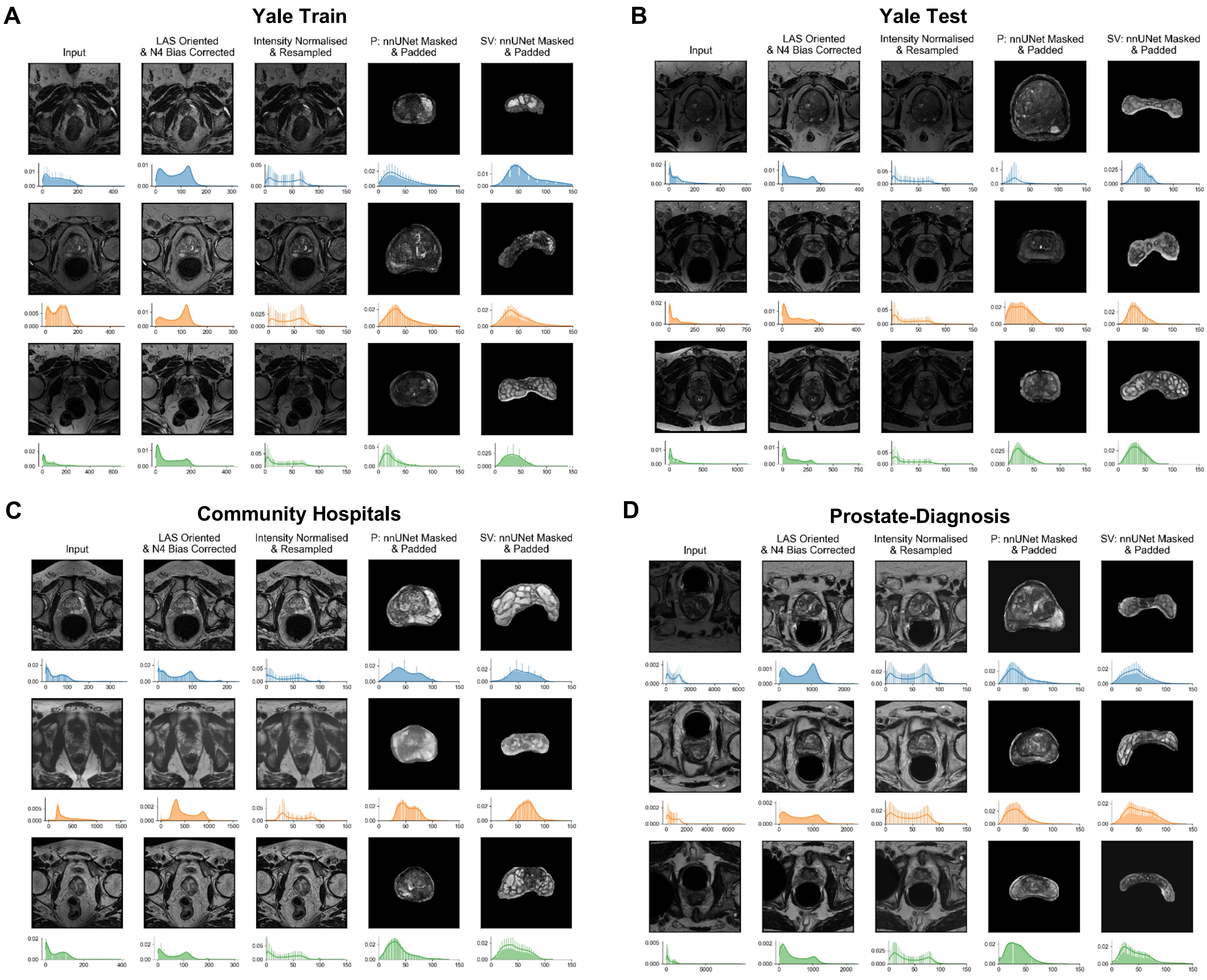


**Supplementary Figure 2 I Example Images and Probability Distribution Functions for Preprocessing Steps Across Cohorts. a** Example images and histograms across Yale train set, **b** examples across Yale test set, **c** examples across community hospitals, **d** examples across public dataset. Abbreviations: **P:** Whole Prostate Gland **SV:** Seminal Vesicles.

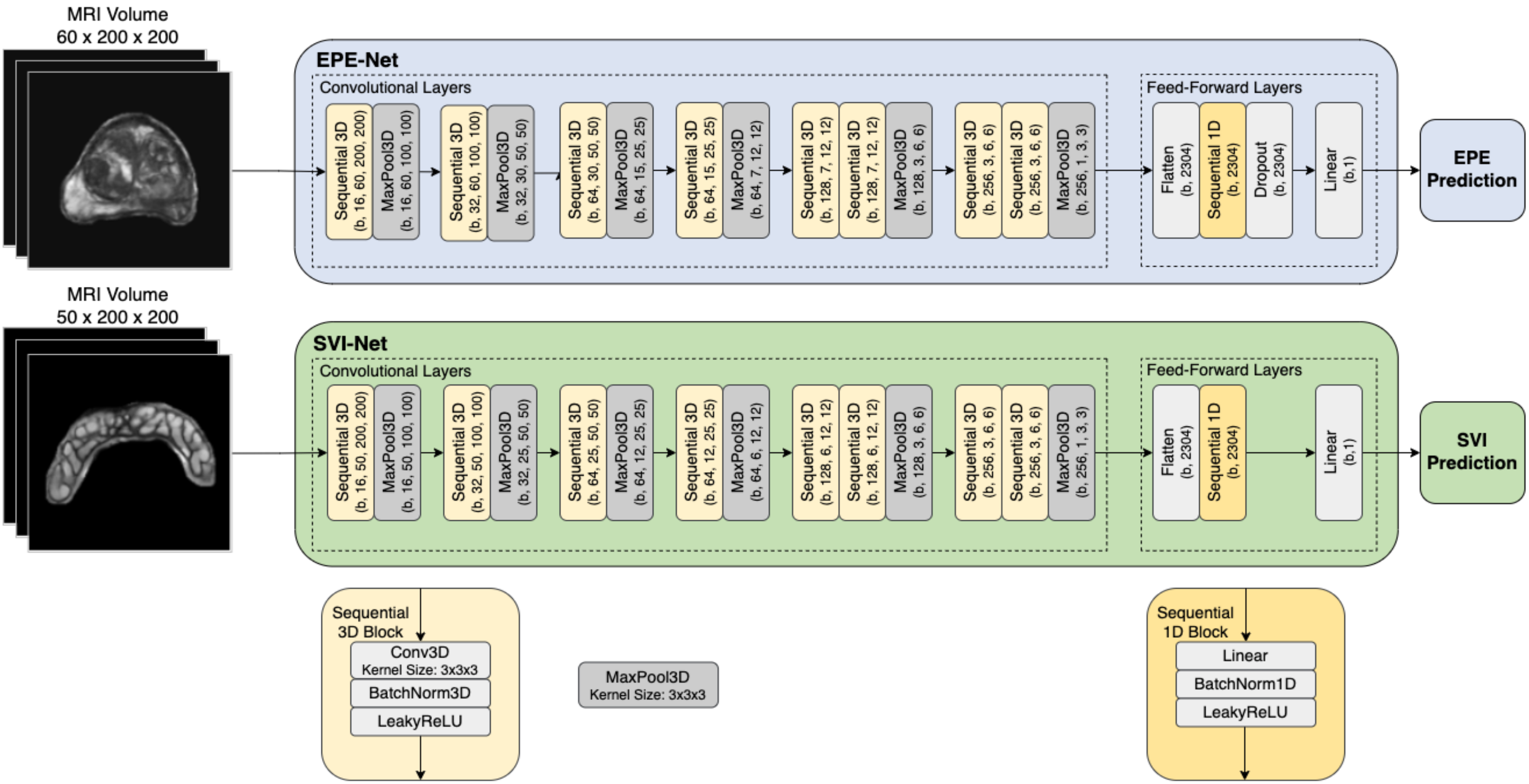


**Supplementary Figure 3 | Deep Learning Architectures for the EPE and SVI Models.** The inputs to the networks are preprocessed T2W axial oriented MRI volumes of either segmented whole prostate glands or seminal vesicles for the EPE and SVI models, respectively. The input sizes are 60 x 200 x 200 for the EPE model and 50 x 200 x 200 for the SVI model. The convolutional layers represent eight layers of convolutions, which are performed on inputs to extract 2304 imaging features. The resulting matrices from the output of the last convolutional layer are first flattened to derive a vector of features. The features are then passed through the feed-forward network layers to make a binary prediction of either EPE or SVI, with sigmoid activation of the final logits. For the EPE model only, the feature vectors are also passed through a dropout layer prior to performing a binary classification. The basic building blocks for these networks consist of a Sequential3D block, Sequential1D block, and MaxPool3D block. The Sequential3D block consists of a 3D Convolutional Layer, a 3D Batch Normalization Layer, and a Leaky Rectified Linear Unit Layer for activation. The Sequential1D block consists of a Densely Connected Linear Layer, a 1D Batch Normalization Layer, and a Leaky Rectified Linear Unit Layer. Kernel sizes for all convolutional and max pooling layers are 3 x 3 x 3. Abbreviations: **MRI:** Magnetic Resonance Imaging, **T2W:** T2 Weighted, **EPE:** Extra-prostatic Extension, **SVI:** Seminal Vesicle Invasion, **b**: Batch-Size.

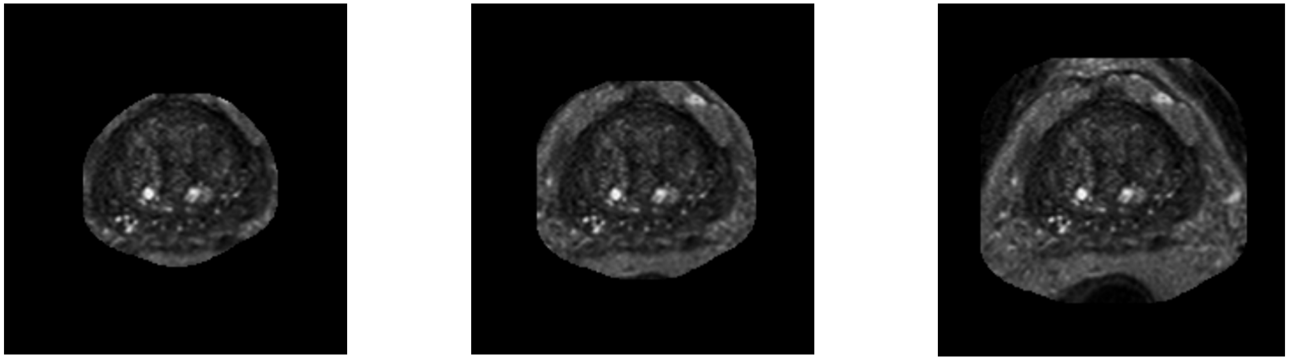

Dilation Kernel: (6, 6, 6) Dilation Kernel: (25, 25, 25) Dilation Kernel: (50, 50, 50)

**Supplementary Figure 4 | Example Images of Prostate Volumes with Expanded Mask Dilation Kernels for the EPE Model Experiments.** Dilation kernels are used to expand and smoothen the whole prostate gland segmentation mask derived from nnU-Net. Three different kernel sizes for dilating segmentations were tested to explore the effect of increasing extracapsular imaging context on the performance of the model. The baseline kernel size for the EPE model was (6, 6, 6). Kernel sizes of (25, 25, 25) and (50, 50, 50) were also tested.

**Supplementary Table 2. The EPE Model Performance Metrics Across Experimental Modifications to Architecture**

Performance metrics are calculated over the Yale test set for each experimental change in EPE-Net architecture

95% confidence intervals are calculated with ten thousand iterations of bootstrap resampling of the Yale test set

| Experiment | Mask Dilation Kernel | Convolutional Layers | Skip Connections | AUC (95%CI) | ACC (95% CI) | SN (95% CI) | SP (95% CI) | NPV (95% CI) | PPV (95% CI) | F1-Score (95% CI) |
|---|---|---|---|---|---|---|---|---|---|---|
| Baseline (EPE-Net) | (6, 6, 6) | 8 | - | 0.713 (0.597 - 0.824) | 0.654 (0.556 - 0.753) | 0.65 (0.500 - 0.795) | 0.659 (0.513 - 0.800) | 0.659 (0.511 - 0.800) | 0.65 (0.500 - 0.795) | 0.65 (0.519 - 0.762) |
| Increased Mask Size | (25, 25, 25) | 8 | - | 0.642 (0.515 - 0.761) | 0.642 (0.531 - 0.741) | 0.7 (0.553 - 0.838) | 0.585 (0.432 - 0.732) | 0.667 (0.500 - 0.821) | 0.622 (0.475 - 0.762) | 0.659 (0.528 - 0.766) |
| | (50, 50, 50) | 8 | - | 0.666 (0.544 - 0.779) | 0.63 (0.519 - 0.728) | 0.5 (0.342 - 0.650) | 0.756 (0.619 - 0.881) | 0.608 (0.469 - 0.740) | 0.667 (0.485 - 0.829) | 0.571 (0.415 - 0.697) |
| Fewer Layers | (6, 6, 6) | 6 | - | 0.667 (0.546 - 0.780) | 0.58 (0.469 - 0.691) | 0.55 (0.393 - 0.705) | 0.61 (0.457 - 0.757) | 0.581 (0.429 - 0.730) | 0.579 (0.417 - 0.735) | 0.564 (0.423 - 0.689) |
| Increased Layers | (6, 6, 6) | 10 | - | 0.713 (0.594 - 0.821) | 0.642 (0.531 - 0.741) | 0.5 (0.345 - 0.659) | 0.78 (0.646 - 0.900) | 0.615 (0.481 - 0.750) | 0.69 (0.500 - 0.852) | 0.58 (0.424 - 0.709) |
| Skip Connections | (6, 6, 6) | 10 | + | 0.714 (0.593 - 0.823) | 0.679 (0.580 - 0.778) | 0.75 (0.607 - 0.880) | 0.61 (0.459 - 0.756) | 0.714 (0.558 - 0.857) | 0.652 (0.511 - 0.786) | 0.698 (0.575 - 0.800) |
| ResNet18 | (6, 6, 6) | 18 | + | 0.656 (0.530 - 0.768) | 0.593 (0.481 - 0.704) | 0.625 (0.472 - 0.771) | 0.561 (0.405 - 0.714) | 0.605 (0.444 - 0.758) | 0.581 (0.429 - 0.729) | 0.602 (0.464 - 0.717) |

Abbreviations: **EPE:** Extraprostatic Extension, **AUC:** Area Under Receiver Operating Characteristic Curve, **ACC:** Accuracy, **SN:** Sensitivity or Recall, **SP:** Specificity, **NPV:** Negative Predictive Value, **PPV:** Positive Predictive Value or Precision, **CI:** Confidence Interval

**Supplementary Table 3. The SVI Model Performance Metrics Across Training Sequences**

Performance metrics are calculated over the Yale test set at the end of each training sequence for SVI-Net

95% confidence intervals are calculated with ten thousand iterations of bootstrap resampling of the Yale test set

| Experiment | SVI- to SVI+ Training Ratio | AUC (95%CI) | ACC (95% CI) | SN (95% CI) | SP (95% CI) | NPV (95% CI) | PPV (95% CI) | F1-Score (95% CI) |
|---|---|---|---|---|---|---|---|---|
| Sequence 1 | 1:1 | 0.67<br>(0.603 - 0.826) | 0.85<br>(0.804 - 0.898) | 0.44<br>(0.250 - 0.636) | 0.9<br>(0.858 - 0.939) | 0.931<br>(0.894 - 0.964) | 0.344<br>(0.184 - 0.519) | 0.386<br>(0.218 - 0.540) |
| Sequence 2 | 1:3 | 0.693<br>(0.573 - 0.813) | 0.766<br>(0.711 - 0.821) | 0.6<br>(0.400 - 0.793) | 0.786<br>(0.730 - 0.840) | 0.943<br>(0.906 - 0.976) | 0.25<br>(0.143 - 0.362) | 0.353<br>(0.217 - 0.477) |
| Sequence 3 | 1:6 | 0.717<br>(0.599 - 0.837) | 0.787<br>(0.732 - 0.838) | 0.64<br>(0.450 - 0.833) | 0.805<br>(0.749 - 0.856) | 0.949<br>(0.915 - 0.978) | 0.281<br>(0.169 - 0.400) | 0.39<br>(0.250 - 0.516) |
| Sequence 4 | 1:10 | 0.724<br>(0.598 - 0.842) | 0.825<br>(0.774 - 0.872) | 0.64<br>(0.450 - 0.833) | 0.848<br>(0.796 - 0.895) | 0.952<br>(0.919 - 0.979) | 0.333<br>(0.204 - 0.469) | 0.438<br>(0.290 - 0.571) |

Abbreviations: **SVI:** Seminal Vesicle Invasion, **AUC:** Area Under Receiver Operating Characteristic Curve, **ACC:** Accuracy, **SN:** Sensitivity or Recall, **SP:** Specificity, **NPV:** Negative Predictive Value, **PPV:** Positive Predictive Value or Precision, **CI:** Confidence Interval

**Supplementary Table 4.**

**The EPE Model Performance Across Subgroups of Yale Test Cohort**

AUCs were calculated for the indicated patient subgroups across the Yale test cohort
95% confidence intervals calculated with ten thousand iterations of bootstrap resampling

| Category | Subgroup | Sample Size (N) | EPE+ (n) | AUC (95% CI) | |
|---|---|---|---|---|---|
| Age | <65 | 92 | 38 | 0.718 | (0.533 - 0.883) |
| | ≥65 | 116 | 53 | 0.73 | (0.566 - 0.870) |
| Race | White | 173 | 73 | 0.711 | (0.573 - 0.833) |
| | Non-white | 35 | 18 | 0.741 | (0.341 - 0.980) |
| PSA (ng/mL) | ≤10 | 139 | 52 | 0.683 | (0.523 - 0.824) |
| | >10 | 69 | 39 | 0.777 | (0.571 - 0.942) |
| Gleason Score | GS6 & GS7 (3+4) | 98 | 36 | 0.663 | (0.481 - 0.828) |
| | GS7 (4+3) | 49 | 14 | 0.675 | (0.581 - 0.893) |
| | GS8 - GS10 | 60 | 40 | 0.843 | (0.846 - 0.971) |

Abbreviations: **EPE:** Extraprostatic Extension, **AUC:** Area Under Receiver Operating Characteristic Curve, **N:** Number of Patients in Indicated Subgroup, **n:** Number of Positive EPE Cases Among N, **GS:** Gleason Score, **PSA:** Prostate Specific Antigen, **CI:** Confidence Interval

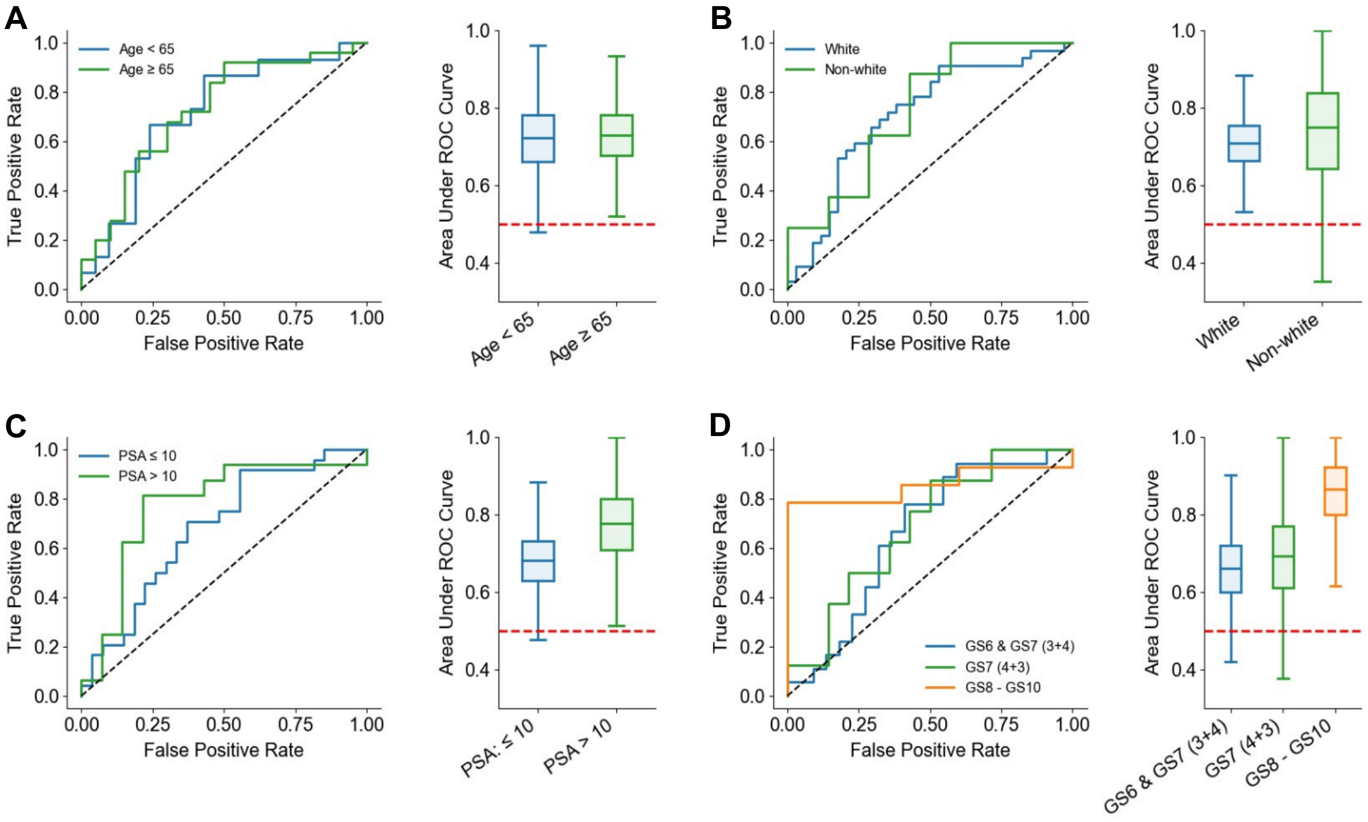


**Supplementary Figure 5 I ROC Curves and AUC Boxplots for the EPE Model Across Subsets of Patients.** ROC curves of the EPE model output for different subsets of patients in the Yale test cohort stratified by **a** age, **b** race, **c** PSA level at diagnosis, and **d** Gleason Scores at biopsy. For ROC curves, the black dashed line indicates no discriminatory ability. Boxplots represent the range of AUCs calculated using ten thousand iterations of bootstrap resampling on the indicated patient subset. The midline of the boxplot represents the median, the upper and lower bounds of the interior box represent the 75th and 25th percentile, respectively, and the upper and lower whiskers represent the complete upper and lower range of AUCs constructed with the bootstrap resampling method. The red dashed line for all boxplots indicates an AUC of 0.50. Abbreviations: **PSA:** Prostate Specific Antigen, **GS:** Gleason Score, **ROC:** Receiver Operating Characteristic, **AUC:** Area Under the ROC Curve.

**Supplementary Table 5.**
**The SVI Model Performance Across Subgroups of Yale Test Cohort**

AUCs were calculated for the indicated patient subgroups across the Yale test cohort
95% confidence intervals calculated with ten thousand iterations of bootstrap resampling
-- Indicates confidence interval could not be calculated due to few positive cases in sample

| Category | Subgroup | Sample Size (N) | SVI+ (n) | AUC (95% CI) | |
|---|---|---|---|---|---|
| Age | <65 | 92 | 10 | 0.769 | (0.526 - 0.947) |
| | ≥65 | 116 | 11 | 0.633 | (0.449 - 0.801) |
| Race | White | 173 | 17 | 0.707 | (0.526 - 0.838) |
| | Non-white | 35 | 4 | 0.76 | (0.635 - 0.938) |
| PSA (ng/mL) | ≤10 | 139 | 11 | 0.596 | (0.393 - 0.793) |
| | >10 | 69 | 10 | 0.811 | (0.611 - 0.960) |
| Gleason Score | GS6 & GS7 (3+4) | 98 | 1 | 0.981 | -- |
| | GS7 (4+3) | 49 | 3 | 0.467 | -- |
| | GS8 - GS10 | 60 | 17 | 0.776 | (0.617 0.909) |

Abbreviations: **SVI:** Seminal Vesicle Invasion, **AUC:** Area Under Receiver Operating Characteristic Curve, **N:** Number of Patients in Indicated Subgroup, **n:** Number of Positive SVI Cases Among N, **GS:** Gleason Score, **PSA:** Prostate Specific Antigen, **CI:** Confidence Interval

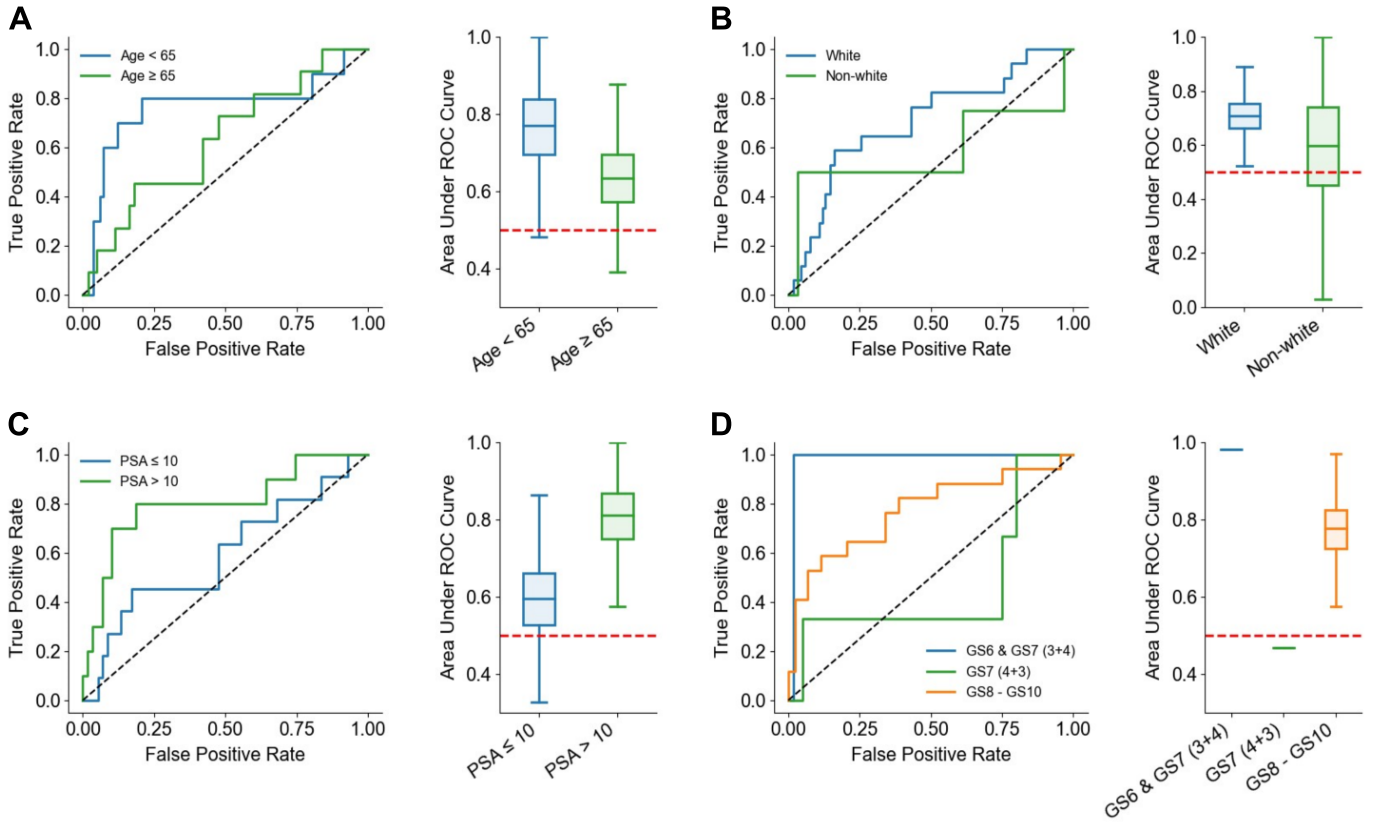


**Supplementary Figure 6 | ROC Curves and AUC Boxplots for the SVI Model Across Subsets of Patients.** ROC curves of the SVI model output for different subsets of patients in the Yale test cohort stratified by **a** age, **b** race, **c** PSA level at diagnosis, and **d** Gleason Scores at biopsy. For ROC curves, the black dashed line indicates no discriminatory ability. Boxplots represent the range of AUCs calculated using ten thousand iterations of bootstrap resampling on the indicated patient subset. The midline of the boxplot represents the median, the upper and lower bounds of the interior box represent the 75th and 25th percentile, respectively, and the upper and lower whiskers represent the complete upper and lower range of AUCs constructed with the bootstrap resampling method. For boxplots of GS less than 8, the single line represents the single AUC of the indicated patient subset of the Yale test set without resampling. The red dashed line for all boxplots indicates an AUC of 0.50. Abbreviations: **PSA:** Prostate Specific Antigen, **GS:** Gleason Score, **ROC:** Receiver Operating Characteristic, **AUC:** Area Under the ROC Curve.

**Supplementary Table 6. Biomarkers, Clinicopathologic, and Clinicopathologic-Imaging Groups Time-dependent AUCs for BCR**

Time-dependent AUCs for predicting BCR across patients included in the indicated subgroup were calculated for each risk group model
95% confidence intervals calculated with ten thousand iterations of bootstrap resai
-- Indicates confidence interval could not be calculated due to few positive cases in sample

| Category | Subgroup | Year | Total (N) | BCR+ (n) | Censored | bEPE | | bSVI | | bI: bEPE + bSVI | | NCCN | | NCCN+bI | | CAPRA | | CAPRA+bI | |
|---|---|---|---|---|---|---|---|---|---|---|---|---|---|---|---|---|---|---|---|
| All Patients | None | 1 | 820 | 88 | 0 | 0.599 | (0.534 - 0.660) | 0.552 | (0.483 - 0.621) | 0.602 | (0.543 - 0.661) | 0.632 | (0.576 - 0.687) | 0.655 | (0.598 - 0.711) | 0.623 | (0.564 - 0.678) | 0.644 | (0.586 - 0.700) |
| | | 3 | 526 | 155 | 294 | 0.562 | (0.505 - 0.617) | 0.571 | (0.517 - 0.626) | 0.589 | (0.540 - 0.639) | 0.587 | (0.537 - 0.636) | 0.615 | (0.565 - 0.666) | 0.606 | (0.555 - 0.656) | 0.627 | (0.578 - 0.677) |
| | | 5 | 339 | 183 | 481 | 0.623 | (0.563 - 0.683) | 0.578 | (0.517 - 0.637) | 0.637 | (0.583 - 0.690) | 0.613 | (0.556 - 0.668) | 0.656 | (0.600 - 0.711) | 0.621 | (0.559 - 0.679) | 0.651 | (0.591 - 0.708) |
| Initial Treatment | Surgery | 1 | 397 | 85 | 0 | 0.636 | (0.565 - 0.705) | 0.558 | (0.481 - 0.632) | 0.631 | (0.567 - 0.694) | 0.702 | (0.646 - 0.758) | 0.723 | (0.666 - 0.778) | 0.725 | (0.666 - 0.783) | 0.743 | (0.684 - 0.798) |
| | | 3 | 313 | 133 | 84 | 0.578 | (0.512 - 0.641) | 0.556 | (0.491 - 0.620) | 0.593 | (0.535 - 0.651) | 0.643 | (0.585 - 0.699) | 0.66 | (0.600 - 0.718) | 0.676 | (0.616 - 0.733) | 0.69 | (0.631 - 0.747) |
| | | 5 | 235 | 153 | 162 | 0.634 | (0.559 - 0.706) | 0.561 | (0.485 - 0.636) | 0.621 | (0.552 - 0.687) | 0.672 | (0.603 - 0.737) | 0.691 | (0.622 - 0.757) | 0.701 | (0.630 - 0.770) | 0.712 | (0.645 - 0.779) |
| | Radiation | 1 | 423 | 3 | 0 | 0.641 | -- | 0.632 | -- | 0.626 | -- | 0.545 | -- | 0.519 | -- | 0.577 | -- | 0.555 | -- |
| | | 3 | 213 | 22 | 210 | 0.642 | (0.495 - 0.777) | 0.696 | (0.579 - 0.801) | 0.666 | (0.547 - 0.779) | 0.573 | (0.431 - 0.711) | 0.626 | (0.492 - 0.755) | 0.637 | (0.505 - 0.760) | 0.673 | (0.543 - 0.786) |
| | | 5 | 104 | 30 | 319 | 0.678 | (0.546 - 0.800) | 0.676 | (0.551 - 0.793) | 0.727 | (0.618 - 0.831) | 0.581 | (0.451 - 0.707) | 0.657 | (0.532 - 0.780) | 0.618 | (0.493 - 0.738) | 0.675 | (0.554 - 0.792) |
| Age (Years) | <65 | 1 | 332 | 48 | 0 | 0.654 | (0.565 - 0.738) | 0.548 | (0.447 - 0.650) | 0.648 | (0.566 - 0.731) | 0.692 | (0.611 - 0.768) | 0.712 | (0.632 - 0.787) | 0.698 | (0.617 - 0.774) | 0.72 | (0.640 - 0.794) |
| | | 3 | 240 | 74 | 92 | 0.641 | (0.560 - 0.718) | 0.578 | (0.495 - 0.658) | 0.659 | (0.586 - 0.728) | 0.646 | (0.572 - 0.717) | 0.687 | (0.615 - 0.756) | 0.679 | (0.606 - 0.749) | 0.711 | (0.639 - 0.778) |
| | | 5 | 170 | 92 | 162 | 0.701 | (0.619 - 0.780) | 0.591 | (0.506 - 0.675) | 0.706 | (0.634 - 0.775) | 0.668 | (0.590 - 0.742) | 0.728 | (0.655 - 0.797) | 0.702 | (0.624 - 0.777) | 0.739 | (0.662 - 0.810) |
| | ≥ 65 | 1 | 488 | 40 | 0 | 0.545 | (0.449 - 0.638) | 0.556 | (0.461 - 0.649) | 0.56 | (0.476 - 0.644) | 0.585 | (0.507 - 0.662) | 0.612 | (0.535 - 0.687) | 0.567 | (0.480 - 0.653) | 0.581 | (0.500 - 0.664) |
| | | 3 | 286 | 81 | 202 | 0.489 | (0.412 - 0.566) | 0.565 | (0.490 - 0.640) | 0.527 | (0.459 - 0.594) | 0.535 | (0.468 - 0.603) | 0.551 | (0.479 - 0.619) | 0.539 | (0.469 - 0.609) | 0.549 | (0.477 - 0.619) |
| | | 5 | 169 | 91 | 319 | 0.536 | (0.446 - 0.626) | 0.564 | (0.476 - 0.651) | 0.567 | (0.488 - 0.645) | 0.556 | (0.473 - 0.636) | 0.578 | (0.494 - 0.661) | 0.535 | (0.449 - 0.622) | 0.552 | (0.464 - 0.636) |
| Race/Ethnicity | White | 1 | 676 | 66 | 0 | 0.582 | (0.507 - 0.657) | 0.558 | (0.485 - 0.631) | 0.59 | (0.522 - 0.656) | 0.642 | (0.579 - 0.705) | 0.661 | (0.601 - 0.721) | 0.636 | (0.572 - 0.699) | 0.653 | (0.591 - 0.713) |
| | | 3 | 441 | 126 | 235 | 0.557 | (0.496 - 0.617) | 0.571 | (0.512 - 0.631) | 0.587 | (0.532 - 0.642) | 0.585 | (0.530 - 0.639) | 0.617 | (0.561 - 0.672) | 0.604 | (0.547 - 0.658) | 0.626 | (0.572 - 0.681) |
| | | 5 | 283 | 147 | 393 | 0.61 | (0.543 - 0.675) | 0.575 | (0.507 - 0.642) | 0.628 | (0.570 - 0.686) | 0.614 | (0.551 - 0.673) | 0.655 | (0.592 - 0.715) | 0.622 | (0.555 - 0.687) | 0.649 | (0.584 - 0.711) |
| | Non-white | 1 | 144 | 22 | 0 | 0.633 | (0.515 - 0.749) | 0.531 | (0.362 - 0.705) | 0.64 | (0.512 - 0.760) | 0.593 | (0.470 - 0.715) | 0.63 | (0.494 - 0.759) | 0.557 | (0.432 - 0.682) | 0.599 | (0.462 - 0.727) |
| | | 3 | 85 | 29 | 59 | 0.574 | (0.438 - 0.707) | 0.57 | (0.426 - 0.710) | 0.594 | (0.472 - 0.713) | 0.596 | (0.477 - 0.710) | 0.605 | (0.473 - 0.731) | 0.599 | (0.477 - 0.720) | 0.624 | (0.498 - 0.746) |
| | | 5 | 56 | 36 | 88 | 0.66 | (0.506 - 0.801) | 0.585 | (0.436 - 0.727) | 0.681 | (0.550 - 0.806) | 0.614 | (0.464 - 0.753) | 0.662 | (0.519 - 0.792) | 0.592 | (0.433 - 0.752) | 0.643 | (0.490 - 0.788) |
| PSA (ng/mL) | ≤ 6 | 1 | 255 | 23 | 0 | 0.623 | (0.479 - 0.755) | 0.624 | (0.480 - 0.759) | 0.657 | (0.537 - 0.770) | 0.7 | (0.581 - 0.806) | 0.737 | (0.621 - 0.840) | 0.653 | (0.533 - 0.762) | 0.698 | (0.578 - 0.806) |
| | | 3 | 165 | 42 | 90 | 0.588 | (0.482 - 0.690) | 0.561 | (0.456 - 0.663) | 0.62 | (0.528 - 0.711) | 0.614 | (0.514 - 0.709) | 0.65 | (0.551 - 0.745) | 0.592 | (0.501 - 0.686) | 0.63 | (0.535 - 0.722) |
| | | 5 | 103 | 50 | 152 | 0.605 | (0.491 - 0.716) | 0.526 | (0.409 - 0.640) | 0.607 | (0.510 - 0.705) | 0.613 | (0.508 - 0.716) | 0.638 | (0.534 - 0.741) | 0.611 | (0.504 - 0.713) | 0.635 | (0.526 - 0.737) |
| | > 6, ≤ 10 | 1 | 276 | 28 | 0 | 0.612 | (0.492 - 0.728) | 0.504 | (0.383 - 0.624) | 0.604 | (0.504 - 0.703) | 0.59 | (0.498 - 0.681) | 0.632 | (0.541 - 0.725) | 0.634 | (0.529 - 0.733) | 0.667 | (0.565 - 0.766) |
| | | 3 | 177 | 51 | 99 | 0.576 | (0.473 - 0.677) | 0.546 | (0.446 - 0.644) | 0.602 | (0.520 - 0.685) | 0.622 | (0.541 - 0.701) | 0.651 | (0.566 - 0.733) | 0.659 | (0.574 - 0.741) | 0.691 | (0.603 - 0.773) |
| | | 5 | 112 | 61 | 164 | 0.614 | (0.509 - 0.715) | 0.501 | (0.393 - 0.611) | 0.627 | (0.535 - 0.717) | 0.628 | (0.520 - 0.727) | 0.67 | (0.571 - 0.763) | 0.656 | (0.555 - 0.756) | 0.692 | (0.594 - 0.781) |
| | > 10, ≤ 20 | 1 | 204 | 24 | 0 | 0.568 | (0.450 - 0.687) | 0.6 | (0.482 - 0.716) | 0.565 | (0.447 - 0.681) | 0.578 | (0.463 - 0.694) | 0.593 | (0.472 - 0.709) | 0.603 | (0.492 - 0.704) | 0.613 | (0.485 - 0.736) |
| | | 3 | 132 | 43 | 72 | 0.479 | (0.372 - 0.585) | 0.608 | (0.505 - 0.710) | 0.524 | (0.422 - 0.625) | 0.456 | (0.359 - 0.555) | 0.494 | (0.384 - 0.597) | 0.561 | (0.464 - 0.652) | 0.555 | (0.451 - 0.655) |
| | | 5 | 91 | 48 | 113 | 0.567 | (0.447 - 0.688) | 0.675 | (0.563 - 0.783) | 0.604 | (0.500 - 0.710) | 0.514 | (0.404 - 0.623) | 0.581 | (0.466 - 0.692) | 0.594 | (0.486 - 0.697) | 0.628 | (0.513 - 0.737) |
| | >20 | 1 | 85 | 13 | 0 | 0.475 | (0.305 - 0.654) | 0.474 | (0.279 - 0.680) | 0.513 | (0.340 - 0.690) | 0.645 | (0.516 - 0.752) | 0.582 | (0.419 - 0.740) | 0.506 | (0.349 - 0.659) | 0.511 | (0.345 - 0.676) |
| | | 3 | 52 | 19 | 33 | 0.562 | (0.384 - 0.731) | 0.584 | (0.412 - 0.748) | 0.573 | (0.414 - 0.727) | 0.648 | (0.525 - 0.765) | 0.627 | (0.472 - 0.771) | 0.565 | (0.399 - 0.726) | 0.589 | (0.416 - 0.756) |
| | | 5 | 33 | 24 | 52 | 0.802 | (0.619 - 0.947) | 0.708 | (0.527 - 0.870) | 0.84 | (0.707 - 0.944) | 0.785 | (0.607 - 0.933) | 0.894 | (0.854 - 0.973) | 0.502 | (0.290 - 0.715) | 0.699 | (0.511 - 0.865) |
| Gleason Score | ≤ GS 7(3+4) | 1 | 364 | 18 | 0 | 0.591 | (0.444 - 0.731) | 0.56 | (0.424 - 0.693) | 0.549 | (0.425 - 0.678) | 0.56 | (0.429 - 0.686) | 0.6 | (0.497 - 0.703) | 0.542 | (0.403 - 0.678) | 0.578 | (0.448 - 0.699) |
| | | 3 | 223 | 44 | 141 | 0.531 | (0.426 - 0.635) | 0.554 | (0.460 - 0.647) | 0.531 | (0.447 - 0.618) | 0.553 | (0.468 - 0.636) | 0.577 | (0.491 - 0.661) | 0.595 | (0.506 - 0.681) | 0.604 | (0.514 - 0.689) |
| | | 5 | 133 | 57 | 231 | 0.576 | (0.473 - 0.676) | 0.517 | (0.415 - 0.616) | 0.569 | (0.486 - 0.655) | 0.581 | (0.491 - 0.668) | 0.615 | (0.523 - 0.704) | 0.676 | (0.587 - 0.762) | 0.678 | (0.586 - 0.763) |
| | GS 7 (4+3) | 1 | 187 | 24 | 0 | 0.586 | (0.464 - 0.706) | 0.545 | (0.393 - 0.701) | 0.598 | (0.475 - 0.715) | 0.473 | (0.417 - 0.543) | 0.561 | (0.443 - 0.678) | 0.48 | (0.365 - 0.599) | 0.537 | (0.417 - 0.655) |
| | | 3 | 114 | 42 | 73 | 0.537 | (0.423 - 0.647) | 0.534 | (0.417 - 0.649) | 0.556 | (0.454 - 0.656) | 0.463 | (0.412 - 0.516) | 0.521 | (0.419 - 0.620) | 0.601 | (0.495 - 0.705) | 0.61 | (0.504 - 0.712) |
| | | 5 | 81 | 45 | 106 | 0.557 | (0.429 - 0.682) | 0.552 | (0.423 - 0.675) | 0.577 | (0.465 - 0.683) | 0.452 | (0.378 - 0.521) | 0.524 | (0.408 - 0.637) | 0.521 | (0.394 - 0.649) | 0.55 | (0.425 - 0.674) |
| | GS 8 - GS 10 | 1 | 269 | 46 | 0 | 0.536 | (0.446 - 0.625) | 0.524 | (0.431 - 0.618) | 0.559 | (0.478 - 0.643) | 0.548 | (0.473 - 0.626) | 0.579 | (0.497 - 0.660) | 0.514 | (0.420 - 0.606) | 0.538 | (0.445 - 0.630) |
| | | 3 | 189 | 69 | 80 | 0.549 | (0.463 - 0.635) | 0.591 | (0.506 - 0.676) | 0.608 | (0.529 - 0.685) | 0.56 | (0.491 - 0.630) | 0.623 | (0.545 - 0.699) | 0.471 | (0.383 - 0.558) | 0.527 | (0.442 - 0.613) |
| | | 5 | 125 | 81 | 144 | 0.653 | (0.548 - 0.752) | 0.634 | (0.533 - 0.733) | 0.695 | (0.603 - 0.781) | 0.628 | (0.557 - 0.696) | 0.724 | (0.634 - 0.808) | 0.511 | (0.415 - 0.611) | 0.598 | (0.500 - 0.695) |

Abbreviations: **BCR:** Biochemical Reccurrence, **AUC:** Area Under Receiver Operating Characteristic Curve, **bEPE:** Bioimaging Marker for Extraprostatic Extension, **bSVI:** Bioimaging Marker for Seminal Vesicle Invasion, **bI:** Combined Bioimaging Marker of EPE and SVI, **NCCN:** National Comprehensive Cancer Network Risk Groups, **CAPRA:** Cancer of the Prostate Risk Assessment Risk Groups, **NCCN+bI:** Combined NCCN and bI Clinicopathologic-Imaging Risk Groups, **CAPRA+bI:** Combined CAPRA and bI Clinicopathologic-Imaging Risk Groups, **PSA:** Prostate Specific Antigen, **GS:** Gleason Score, **N:** Total Number of Cases for the Indicated Subgroup and Timepoint, **n:** Number of Positive BCR Cases Among N, **CI:** Confidence Interval

**Supplementary Table 7. Performance of Biomarkers, Clinicopathologic, and Combined Clinicopathologic-Imaging Risk Groups In Predicting Adverse Pathologies in Post Surgical Specimens**

AUCs calculated represent each models discriminatory performance in predicting the indicated pathological outcome among the subset of patients who had complete pathological annotations after treatment with Radical Prostatectomy (N = 387)
95% confidence intervals calculated with ten thousand iterations of bootstrap resampling
*n* represents the number of cases positive for the indicated pathological outcome among N

| Model | PSM, AUC (95% CI) *n* = 167 | | pEPE, AUC (95% CI) *n* = 202 | | pSVI, AUC (95% CI) *n* = 58 | | pEPE and pSVI, AUC (95% CI), *n* = 53 | |
|---|---|---|---|---|---|---|---|---|
| bSVI | 0.575 | (0.516 - 0.633) | 0.571 | (0.512 - 0.628) | 0.699 | (0.615 - 0.777) | 0.69 | (0.602 - 0.774) |
| bEPE | 0.549 | (0.490 - 0.607) | 0.666 | (0.610 - 0.719) | 0.624 | (0.539 - 0.704) | 0.644 | (0.554 - 0.728) |
| bI: bEPE + bSVI | 0.567 | (0.516 - 0.619) | 0.661 | (0.612 - 0.708) | 0.676 | (0.600 - 0.750) | 0.691 | (0.610 - 0.766) |
| NCCN | 0.501 | (0.445 - 0.556) | 0.658 | (0.605 - 0.708) | 0.737 | (0.665 - 0.801) | 0.756 | (0.686 - 0.821) |
| NCCN+bI | 0.535 | (0.479 - 0.591) | 0.708 | (0.658 - 0.756) | 0.767 | (0.693 - 0.832) | 0.787 | (0.716 - 0.851) |
| CAPRA | 0.529 | (0.472 - 0.586) | 0.625 | (0.568 - 0.679) | 0.679 | (0.603 - 0.751) | 0.71 | (0.636 - 0.779) |
| CAPRA+bI | 0.547 | (0.490 - 0.603) | 0.666 | (0.612 - 0.718) | 0.723 | (0.650 - 0.792) | 0.765 | (0.693 - 0.818) |

Abbreviations: **AUC:** Area Under Receiver Operating Characteristic Curve, **PSM:** Positive Surgical Margins, **pEPE:** Pathological Extraprostatic Extension, **pSVI:** Pathological Seminal Vesicle Invasion, **bEPE:** Bioimaging Marker for Extraprostatic Extension, **bSVI:** Bioimaging Marker for Seminal Vesicle Invasion, **bI:** Combined Bioimaging Marker of EPE and SVI, **NCCN:** National Comprehensive Cancer Network Risk Groups, **CAPRA:** Cancer of the Prostate Risk Assessment Risk Groups, **NCCN+bI:** Combined NCCN and bI Clinicopathologic-Imaging Risk Groups, **CAPRA+bI:** Combined CAPRA and bI Clinicopathologic-Imaging Risk Groups, **CI:** Confidence Interval

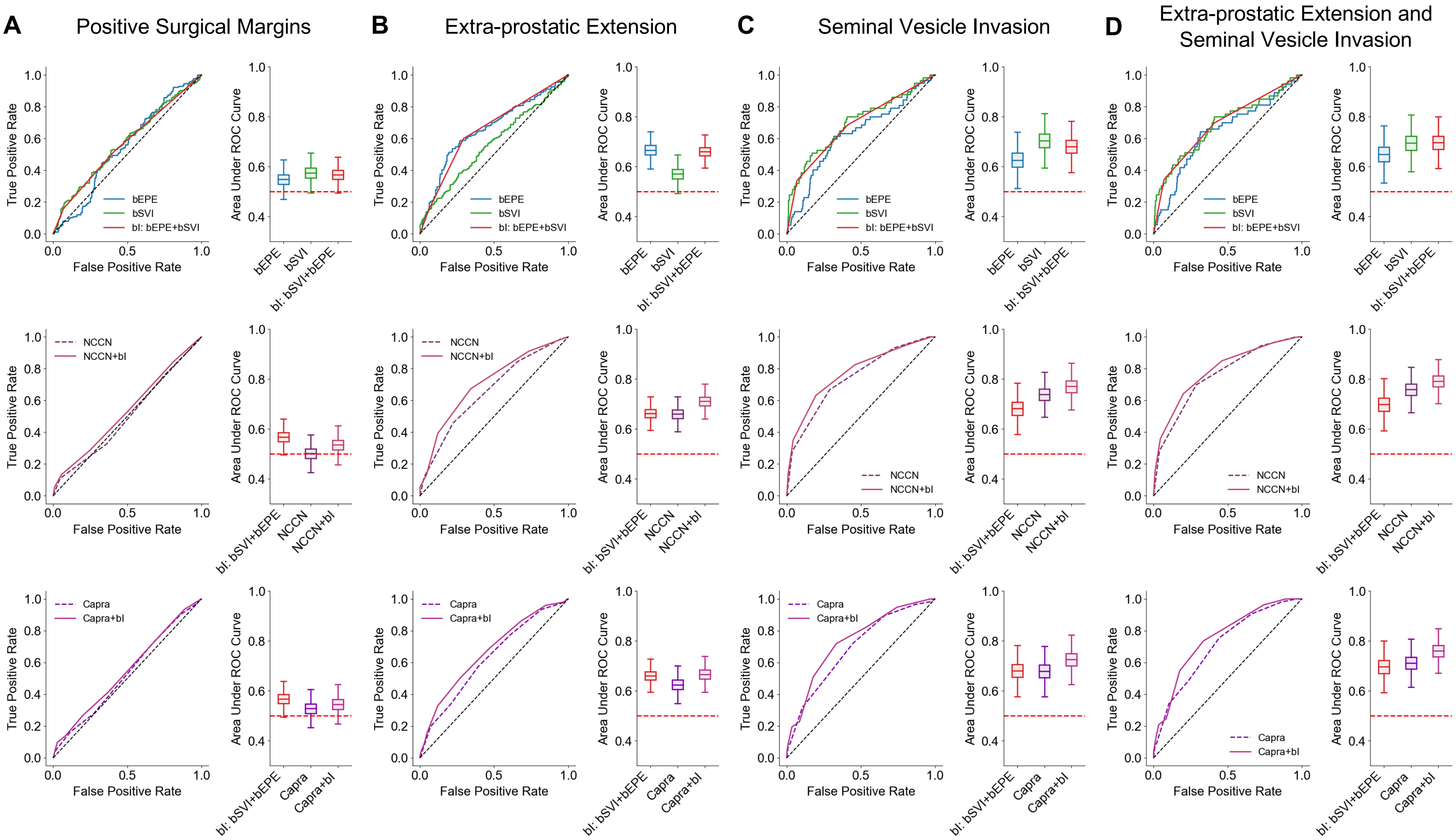


**Supplementary Figure 7 | ROC Curves and AUC Boxplots for Biomarkers, Clinicopathologic and Clinicopathologic-Imaging Risk Groups in Predicting Adverse Pathologies.** ROC curves and Boxplots of AUCs for predicting **a** Positive Surgical Margins, **b** Extra-prostatic Extension, **c** Seminal Vesicle Invasion, and **d** both Extra-prostatic Extension and Seminal Vesicle Invasion. For each panel of adverse pathologies, the first row shows the ROCs and AUCs of the imaging biomarkers bEPE, bSVI, and bI. The second row compares the clinicopathologic NCCN risk groups with the combined clinicopathologic-imaging risk groups of NCCN+bI. The third row compares the clinicopathologic CAPRA risk groups with the combined clinicopathologic-imaging risk groups of CAPRA+bI. For ROC curves, the black dashed line indicates no discriminatory ability. All boxplots represent the range of AUCs calculated using ten thousand iterations of bootstrap resampling. The midline of the boxplot represents the median, the upper and lower bounds of the interior box represent the 75th and 25th percentile, respectively, and the upper and lower whiskers represent the complete upper and lower range of AUCs constructed with the bootstrap resampling method. The red dashed line for all boxplots indicates an AUC of 0.50. Abbreviations: **ROC:** Receiver Operating Characteristic, **AUC:** Area Under ROC Curve, **bEPE:** Bioimaging Marker for Extra-prostatic Extension, **bSVI:** Bioimaging Marker for Seminal Vesicle Invasion, **bI:** Combined Bioimaging Marker of EPE and SVI, **NCCN**: National Comprehensive Cancer Network Risk Groups, **CAPRA:** Cancer of the Prostate Risk Assessment Risk Groups, **NCCN+bI:** Combined NCCN and bI Clinicopathologic-Imaging Risk Groups, **CAPRA+bI:** Combined CAPRA and bI Clinicopathologic-Imaging Risk Groups

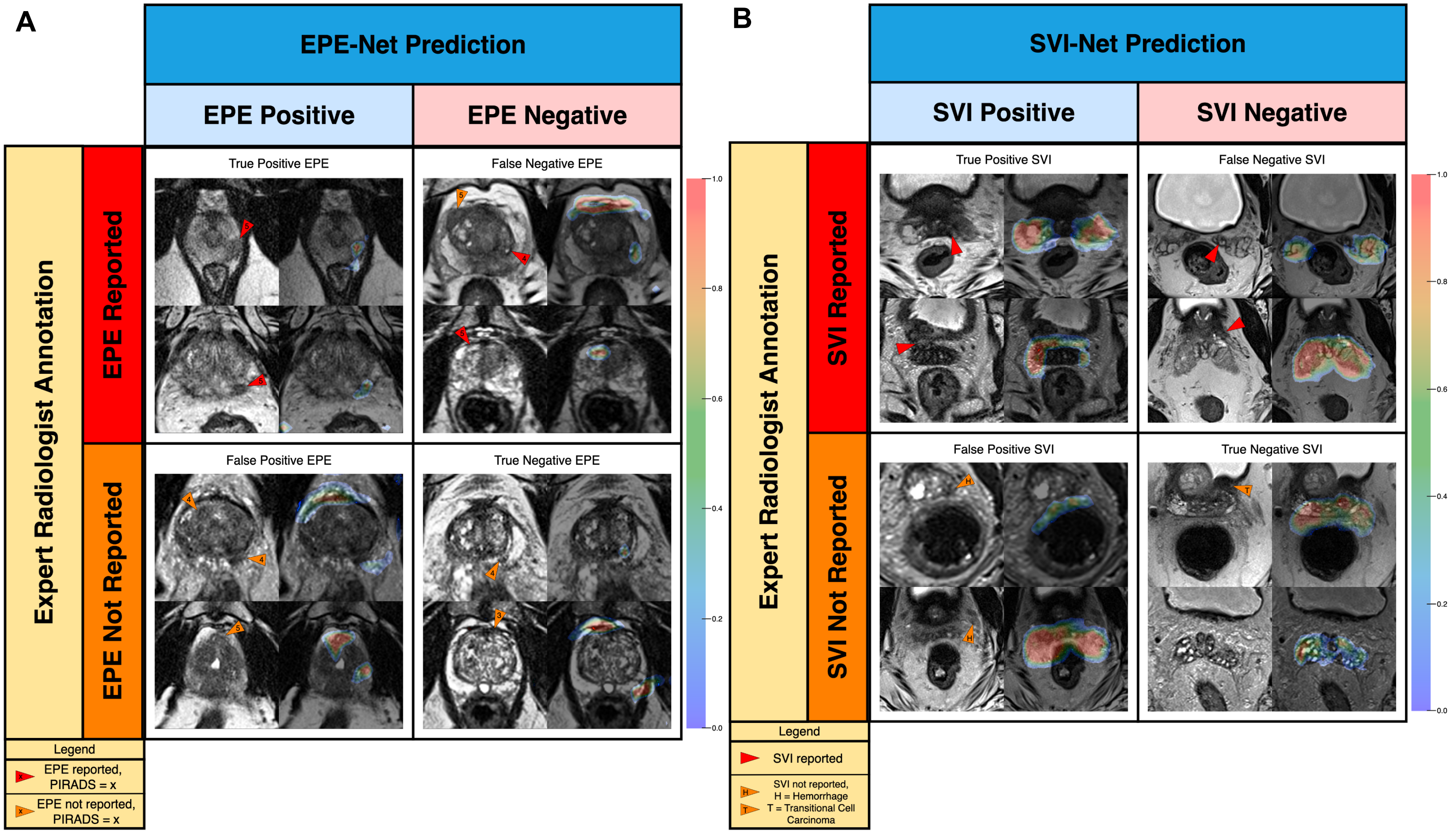


**Supplementary Figure 8 l Representative Gradient-Weighted Class Activation Maps (Grad-CAMs) for the EPE and SVI Models.** Representative Grad-CAMs for correctly and incorrectly classified images for both the EPE (a) and SVI (b) models. Arrows represent diagnostic radiologist findings.
